# Maintaining community-based cohorts in sub-Saharan Africa: An evaluation of participant attrition in SchistoTrack

**DOI:** 10.1101/2024.10.01.24314711

**Authors:** Christin Puthur, Betty Nabatte, Benjamin Tinkitina, Narcis B. Kabatereine, Goylette F. Chami

## Abstract

**Background:** Understanding participant attrition in longitudinal studies is essential for maintaining cohorts, establishing targeted interventions, and assessing potential biases introduced in study analyses. Yet, limited metrics, models, and long-term assessments exist for the evaluation of community-based cohorts in sub-Saharan Africa.

**Methods:** We prospectively assessed participant attrition in the SchistoTrack cohort. A total of 2844 individuals aged 5-92 years were examined from 1445 randomly sampled households across three rural Ugandan districts. Baseline data on sociodemographics, medical history, spatial factors, and clinical examinations were collected in 2022, with annual and seasonal follow-ups analysed to 2024. Profiles of attriters and rejoiners were established with logistic regressions, while the timing of the first attrition event was analysed in multinomial models. Annual community engagement was conducted.

**Findings:** Overall attrition rates were stable across the years ranging from 21-24.8%. Attriter profiles were established within the first year, with only borderline significant factors identified. Home ownership, compared to renting was negatively related to attrition (0.773; CI 0.599–0.998). And, each additional household member reduced the likelihood of attrition (0.923; CI 0.863–0.987). Higher education was positively associated with attrition (1.077; CI 1.047–1.108). Fishermen were not more likely than other individuals to have an attrition event, either overall or seasonally. 40.1% (240/598) of participants who dropped out from the first major follow-up rejoined the study at the following timepoint. Schistosome infection and the need for schistosomiasis-related medical referrals were not associated with later attrition when compared to uninfected individuals and individuals with referrals for ancillary causes or no needed referral. Communicating clinical findings and adjusting incentives across the years did not negatively impact study participation.

**Interpretation:** By providing metrics and models for tracking attrition, our attrition analysis framework can guide the design and evaluation of community-based cohorts in rural sub-Saharan Africa.

**What is already known on this topic:** Participant attrition in longitudinal studies is common and, if not measured and accounted for, can lead to analytical biases and reduced statistical power to produce substandard study designs as well as reduced access to continued care for participants needing further treatment.

**What this study adds:** We comprehensively tracked attrition in a large-scale prospective cohort (SchistoTrack). Attrition at the levels of the individuals, households, villages, and districts was examined in rural Uganda. We investigated a wide range of biomedical, social, spatial, and cultural factors, and developed generalisable procedures and metrics for examining attrition both temporally and seasonally in community-based studies in sub-Saharan Africa.

**How this study might affect research, practice or policy:** More cohort studies urgently are needed in sub-Saharan African countries to understand disease development within these diverse populations. We provide a comprehensive framework to monitor and evaluate the impact of attrition to promote the successful maintenance of rigorous cohort studies. The attrition rates established here can also be used more widely to design effective participant sampling and sample size calculations across different epidemiological study designs.

## Background

Longitudinal studies (cohorts) in epidemiology are essential for the identification of key risk factors of disease without reverse causality and to understand disease progression. Participant recruitment and retention underpin the success of any longitudinal study trying to ascertain individual or population-level trends. Three main ways in which attrition could statistically affect results from a longitudinal study include reduced statistical power, reduced generalisability, and selection bias ^1^. High attrition can lead to reduced analytical power and the ability to detect small effects. Non-random attrition reduces the generalisability of study findings by potentially compromising the initial representativeness of the study population. Participant characteristics associated with both attrition and study outcomes may lead to selection biases in the estimates of outcomes ^2 3^. Importantly, high rates of attrition may be an indicator of ineffective community engagement or diminished trust and receptiveness of study participants ^4^. At worst, attrition could impact participant care needs including treatment and medical referrals directly provided by a study or as a result of study coordination with the local health system ^5^. Yet, attrition in community-based cohort studies in sub-Saharan Africa, especially studies focused on prospective analyses of individual-based outcomes, remains poorly understood despite the urgent need for more cohort studies in these areas ^67^.

In sub-Saharan Africa, the study of attrition has been focused on non-adherence or loss to followup (LTFU) in specialist populations to monitor the treatment of tuberculosis (TB) patients ^8^, or antiretroviral treatment (ART) administration for people living with HIV (PLHIV). A systematic review and meta-analyses of risk factors associated with LTFU and non-adherence to ART in low and middle-income countries ^9^, which included 87.5% of studies from sub-Saharan Africa, found that the percentage of patients identified as LTFU varied widely, ranging from 2.8% to 65.6%. This wide range poses challenges for sample size estimations when designing cohorts. Most studies in the review had a follow-up period of 6 to 24 months with LTFU defined as being away from care for a minimum period ranging from 30 to 90 days. Studies with multiple follow-up timepoints only reported annual attrition rates and did not consider participants who might have rejoined after missing a follow-up.

A wide range of participant characteristics that vary across studies have been associated with an increased likelihood of LTFU for ART adherence ^9^. These characteristics included male sex, younger age, lower educational attainment, being single, unemployment, advanced clinical stage, low weight, poor functional status, poor ART adherence, and participant nondisclosure of HIV-positive status. The LTFU characteristics have been used during ART initiation to screen participants likely to be LTFU for targeted interventions. It remains an open question as to whether such characteristics are generalisable beyond specialist populations of PLHIV. In contrast, in fields with possibly fewer financial resources for follow-up such as malaria, migration was a major reason for attrition ^10^. Apart from studies on TB and HIV, participants who do not show up at initial follow-ups are often excluded from later timepoints of cohort studies ^11^. Community-based studies in sub-Saharan Africa often lack an assessment of factors statistically associated with attrition unless the goal of the study is to assess a population for potential vaccine trials ^12^. It remains an open question as to how best to model attrition to account for study-specific factors such as the length of study, type of attrition, retention methods and candidate predictors of attrition.

The aim of this study was to model attrition and rejoining across diverse populations in rural Uganda, focusing on a large-scale prospective, community-based cohort – SchistoTrack. We conducted a comprehensive analysis of attrition over three years for 2844 individuals aged five years and older from 38 villages of three districts across Western and Eastern Uganda. We addressed the following question. What is the relative influence of medical history, and sociodemographic and spatial factors on attrition across and within study timepoints?

## Materials and Methods

### Study design and participant sampling

This study was nested within SchistoTrack ^13^. This ongoing cohort was established in 2022 in three rural districts of Pakwach, Buliisa, and Mayuge along the River Nile, Lake Albert, and Lake Victoria in Uganda, respectively. SchistoTrack was set up as a community-based prospective cohort with annual follow-ups to study co-infection interactions and concurrent long-term health conditions associated with schistosomiasis.

In January 2022, 1459 households were randomly selected with oversampling from 38 rural villages. Within each household, the head and/or their spouse selected one adult (18+ years) and one child (5+ years) to participate in a clinical study. A total of 2885 individuals were recruited with consent for the baseline examination in 2022, where treatment with praziquantel (PZQ) tablets was provided at each annual study visit irrespective of schistosomiasis diagnosis. The start of the cohort was conducted after a period of extreme flooding which displaced entire villages in the study districts where high water levels remained during study visits. Additionally, the cohort study commenced after the COVID-19 lockdown restrictions were lifted, though several public health measures remained in place in 2022. These measures included nightly curfews from 7 pm to 7 am, mandatory testing for travel, and requirements to wear masks. Moreover, schools were only just beginning to reopen during this time. A short-term drug efficacy follow-up was conducted 3-4 weeks later (March 2022) to evaluate the efficacy of the PZQ treatment on infection status and intensity. Annual follow-up examinations were carried out in January to February 2023 (Jan-Feb 2023), and January to February 2024 (Jan-Feb 2024). Both of these timepoints represent the dry seasons in the study districts with the Jan-Feb 2024 timepoint experiencing longer rains from the preceding rainy season. Additionally, a follow-up examination was conducted in October 2023 (Oct 2023) to assess the seasonal variation (here the rainy season) in schistosome and malaria infections. Two cohort expansions were conducted in 2023 with 14 new villages added and in 2024 with more households added from previously engaged study villages.

Our analysis explores the participation of 2885 individuals from the baseline examination, but does not consider new participants from cohort expansion. As the aim of this study was to investigate modifiable factors that could affect study participation, deaths were excluded from the analysis. Of the 2885 participants in the baseline examinations in Jan-Feb 2022, 41 deaths were recorded leaving 2844 individuals in the final analysis. The participant flow diagram is shown in Figure 1.

**Fig. 1.**
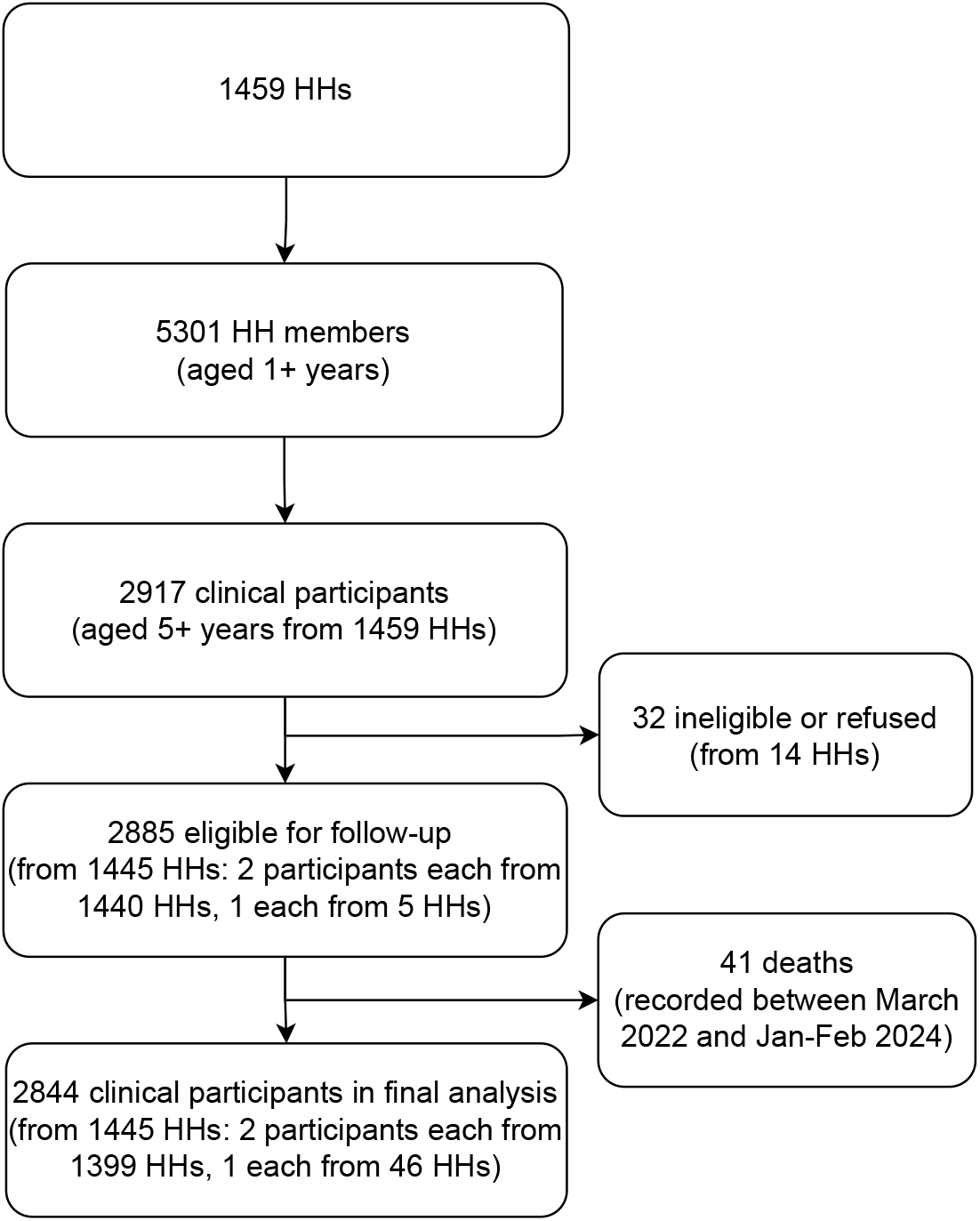
Participant flow diagram.

### Data collection

Data collection was facilitated through digital surveys created using Open Data Kit (ODK) Collect v2022.4.2, and ODK Collect v2023.2.4. Data entry through ODK surveys was done using Android tablets with the software version Android 9.0 Pie. Sociodemographic information and medical history of all members aged one year and above of the sampled households were collected by surveyors at the time of recruitment. Surveyors provided containers for stool and urine samples to village health team members (VHTs) for distribution to the household participants selected for the clinical study. After participants in a study village were mobilised by VHTs and local village leaders, study teams spent two days for each village to conduct clinical examinations for those selected residents. Clinical stations were set up at a central location that could serve/border two villages such as a school or church.

### Community engagement and health outreach

Community engagement was conducted before every timepoint. District sensitisation and mobilisation of participants were led by district focal people, community leaders, and VHTs through community meetings. During the baseline examinations in 2022, all adult participants received 10,000 Ugandan Shillings (approx. $3) as compensation, while children were given pens and notebooks. At the follow-up timepoints, large household-sized soap bars were provided in place of cash as incentives to each household. Participants were informed about the type of compensation during mobilisation. Participants were also informed of their schistosome infection status from their last visit.

Technicians, nurses, and sonographers examined participants. Clinical stations included separate areas for stool and urine sample drop-off, blood tests, nurse anthropometry, nurse palpation, ultrasonography, and food, drink, PZQ and antimalarial treatment. The treatment was provided by local government nurses as part of the study outreach activities. Participants in need of further medical attention were provided medical referrals, and transported to local facilities for serious cases. The study coordinated with local doctors, followed up the care of participants, and where needed financially supported direct costs of additional diagnostic tests that were not available in government health facilities.

### Outcomes

Our attrition outcomes focus on three major study timepoints including the first annual follow-up of Jan-Feb 2023, the seasonal timepoint of Oct 2023, and the second annual follow-up of Jan-Feb 2024. Reasons for attrition were communicated to the study team by VHTs. These reasons were broadly characterised to capture different factors such as migration, sickness, work commitments, school attendance, or refusal to participate. The VHTs obtained these reasons during participant mobilisation and follow-up during examination days.

To identify factors associated with attrition, participants were classified as non-attrited if they attended all timepoints under consideration and as attrited if they missed one or more study timepoints. The attrition status was recorded based on observation by the study teams, noting whether participants were seen at any clinical stations. This binary outcome also was reconstructed to include the short-term drug-efficacy timepoint of March 2022 as a robustness check for the relevance of short-term attrition. Entire households were classified as non-attrited if at least one participant attended all timepoints.

### Covariates

The history of disease (henceforth referred to as biomedical) and sociodemographic variables were collected using a household survey administered by local surveyors fluent in Lusoga, Lugungu, or Alur and English at the time of baseline recruitment. Spatial variables were defined using waypoint data collected with the household survey as well as waypoints collected of all private drug shops and health centres located within the study catchment in 2022.

Sociodemographic variables of age, sex, tribe, religion, education, and occupation were measured at the individual level. At the household level, variables measured included home quality, social status, household size, number of deaths in the past three years, years of settlement in the village, home ownership, number of rooms in the home, drinking water source, drinking water treatment/safety, and the level of improved sanitation and hygiene facilities. Social status was defined as any adult household member holding or previously having a position in the local council, village health team, religious or clan leadership, or influential beach management committees. Individual-level biomedical factors included presence of any symptoms, history of infectious diseases (IDs), history of non-communicable diseases (NCDs), and history of any disease. To understand whether participant attrition is influenced by the need to care for other household members, binary indicators for the history of IDs and NCDs for all other household members aged one year and older apart from the participant of interest also were constructed. Spatial factors considered in the analysis included household distance (km) to the nearest drug shop and health centre to represent the access to care for the participant outside of the cohort study. For district, Mayuge was set as the reference to look at geographical differences between Western (Pakwach and Buliisa) and Eastern (Mayuge) Uganda.

Detailed definitions of all variables are provided in the Supplementary methods S1.1.

### Statistical analysis

All analyses were compiled in R v4.2.1. Logistic regressions were used for attrition outcomes. For the main outcome of overall attrition, variables were selected by likelihood ratio tests (LRTs) in univariable regressions and included in the adjusted analysis if p-value <0.05. For logistic regression models where the tribe variable was significant, floating absolute risks (FARs) were computed with the Epi package in R ^14^ to test the choice of reference category. To account for the paired sampling design within households, standard errors were clustered at the household level ^15^. Intraclass correlation coefficients (ICCs) for empty multilevel models at the household and village levels were investigated to assess within-cluster and between-cluster variations. Variance inflation factors (VIFs) were computed for covariates from the adjusted model to check for multicollinearity ^16^. The predictive capacity of the model was checked by calculating the mean area under the curve (AUC) of the receiver operating characteristic curve over ten-fold cross-validation ^17^. Keeping the variable selection set after the main outcome model to enable cross-model comparisons, the same model-building procedure was applied to look at attrition outcomes that included the short-term drug efficacy timepoint, were specific to each timepoint, and related to missing a timepoint and later rejoining the study.

To investigate potential attrition bias, binary indicators of periportal fibrosis (PPF) ^18^, schistosomiasis-related referrals as well as exposures of schistosome infection status and a categorical indicator of schistosome infection intensity at each timepoint were examined against attrition at a later timepoint. Chi-squared tests were used with Fisher’s exact test if sample sizes were less than five. Definitions of PPF, medical referrals, and schistosomiasis are provided in Supplementary methods S1.2.

## Results

### Attrition rates

Characteristics of study participants are summarised in Table 1 and Supplementary Tables S1, S2, S3, S4, S5, S6 and S7. Participant flows in and out of the study with the reason for attrition at the follow-up timepoints are presented in the Supplementary Figure S1. The most common reason for non-attendance at each timepoint was inconvenience, such as work commitments, school obligations, or caretaking responsibilities, followed by migrations, with refusals, sickness and other reasons being the least common. For example, of the total number of participants who attrited over the three years, only 1.9% (22/1134) were due to refusals in Jan-Feb 2023. (Frequencies of different attrition reasons at each timepoint are shown in the Supplementary Figure S1.) The overall percentage of participants who attrited from at least one of the three major follow-up timepoints of Jan-Feb 2023, Oct 2023, or Jan-Feb 2024 was 39.9% (1134/2844). Inclusion of the short-term drug-efficacy timepoint of March 2022 negligibly increased the overall rate to 41.3% (1175/2844), which henceforth is not included in summary statistics. There were 13.3% (186/1399) of households where both participants simultaneously missed at least one timepoint across the study. The average attrition rate across all villages was 40.4% (SD: 8.8%, range: 25.3%–56.3%). Attrition in each of the three districts was similar; Pakwach, Buliisa, and Mayuge had attrition of 38.1% (360/944), 43% (407/947), and 38.5% (367/953) respectively.

**Table 1.** Summary statistics of potential predictors of attrition.

| | Total<br>(N=2844) | Attrited<br>(N=1134) | Non-attrited<br>(N=1710) | P-value<br>($\chi^2$/t-test) |
| --- | --- | --- | --- | --- |
| **BIOMEDICAL FACTORS** |  |  |  |  |
| Any symptoms | 129 (4.5%) | 44 (3.9%) | 85 (5.0%) | 0.202 |
| IDs for other HH members | 1739 (61.1%) | 684 (60.3%) | 1055 (61.7%) | 0.484 |
| NCDs for other HH members | 965 (33.9%) | 422 (37.2%) | 543 (31.8%) | 0.003 |
| History of IDs | 1405 (49.4%) | 545 (48.1%) | 860 (50.3%) | 0.259 |
| History of NCDs | 543 (19.1%) | 193 (17.0%) | 350 (20.5%) | 0.025 |
| History of any disease | 1584 (55.7%) | 617 (54.4%) | 967 (56.5%) | 0.277 |
| **SOCIODEMOGRAPHICS** |  |  |  |  |
| Age |  |  |  | <0.001 |
| Mean (SD) | 24.6 (18.0) | 22.6 (16.3) | 25.9 (18.9) |  |
| Sex - Female | 1565 (55.0%) | 631 (55.6%) | 934 (54.6%) | 0.618 |
| Tribe |  |  |  | 0.017 |
| Alur | 1550 (54.5%) | 630 (55.6%) | 920 (53.8%) |  |
| Bagungu | 235 (8.3%) | 108 (9.5%) | 127 (7.4%) |  |
| Musoga | 461 (16.2%) | 187 (16.5%) | 274 (16.0%) |  |
| Other | 598 (21.0%) | 209 (18.4%) | 389 (22.7%) |  |
| Majority religion | 2049 (72.0%) | 808 (71.3%) | 1241 (72.6%) | 0.468 |
| Highest level of education attained |  |  |  | <0.001 |
| Mean (SD) | 3.45 (2.83) | 3.72 (2.86) | 3.28 (2.79) |  |
| Occupation |  |  |  | 0.011 |
| Farmer | 508 (17.9%) | 172 (15.2%) | 336 (19.6%) |  |
| Fisherman | 243 (8.5%) | 98 (8.6%) | 145 (8.5%) |  |
| Fishmonger | 108 (3.8%) | 38 (3.4%) | 70 (4.1%) |  |
| None/Other | 1985 (69.8%) | 826 (72.8%) | 1159 (67.8%) |  |
| Home quality score |  |  |  | 0.209 |
| Mean (SD) | 6.38 (3.56) | 6.48 (3.64) | 6.31 (3.50) |  |
| Household social status<sup>1</sup> | 285 (10.0%) | 101 (8.9%) | 184 (10.8%) | 0.122 |
| Number of individuals in household |  |  |  | 0.001 |
| Mean (SD) | 3.63 (1.38) | 3.53 (1.24) | 3.70 (1.46) |  |
| Deaths in household (past 3 yrs) | 184 (6.5%) | 60 (5.3%) | 124 (7.3%) | 0.045 |
| Years household has lived in village |  |  |  | <0.001 |
| Mean (SD) | 18.9 (14.9) | 17.7 (14.5) | 19.8 (15.2) |  |
| Home owned | 2403 (84.5%) | 929 (81.9%) | 1474 (86.2%) | 0.002 |
| Number of rooms |  |  |  | 0.481 |
| Mean (SD) | 2.17 (1.23) | 2.15 (1.26) | 2.19 (1.21) |  |
| **WATER, SANITATION AND HYGIENE (WASH)** |  |  |  |  |
| Improved drinking water source | 1667 (58.6%) | 656 (57.8%) | 1011 (59.1%) | 0.524 |
| Household treats drinking water | 643 (22.6%) | 251 (22.1%) | 392 (22.9%) | 0.655 |
| Improved sanitation facility in home | 1742 (61.3%) | 672 (59.3%) | 1070 (62.6%) | 0.082 |
| Basic hygiene facility in home | 311 (10.9%) | 114 (10.1%) | 197 (11.5%) | 0.243 |
| **SPATIAL FACTORS** |  |  |  |  |
| Min. dist. (km) to drug shop |  |  |  | 0.092 |
| Mean (SD) | 0.937 (1.15) | 0.892 (1.16) | 0.967 (1.15) |  |
| Min. dist. (km) to gov't. health centre |  |  |  | 0.044 |
| Mean (SD) | 2.30 (1.33) | 2.24 (1.28) | 2.34 (1.36) |  |
| District |  |  |  | 0.057 |
| Buliisa | 947 (33.3%) | 407 (35.9%) | 540 (31.6%) |  |
| Mayuge | 953 (33.5%) | 367 (32.4%) | 586 (34.3%) |  |
| Pakwach | 944 (33.2%) | 360 (31.7%) | 584 (34.2%) |  |

<sup>1</sup> Household social status indicates whether one or more members of the participant's household held a position in the local administration.

Attrition in Jan-Feb 2023, Oct 2023, and Jan-Feb 2024 was 21% (598/2844), 23.9% (681/2844), and 24.8% (706/2844) respectively (Supplementary Figure S2). Most participants had their first attrition in Jan-Feb 2023. The percentage of participants who had their first attrition event in JanFeb 2023, Oct 2023, and Jan-Feb 2024 was 21% (598/2844), 11.4% (323/2844), and 7.5% (213/2844) respectively. The household attrition rate, referring to instances where both study participants from a household missed the same timepoint, was 8.1% (113/1399) in Jan-Feb 2023, 8.7% (122/1399) in Oct 2023, and 9.4% (131/1399) in Jan-Feb 2024. The average attrition rate across all villages was similar across timepoints with similar variability. There was 21% attrition (SD: 7.2%, range: 6.2%–33.3%) for Jan-Feb 2023, 23.9% (SD: 7.2%, range: 9.9%–41.2%) for Oct 2023, and 24.8% (SD: 7.4%, range: 10.1%–41%) for Jan-Feb 2024. Attrition was lowest in Pakwach as compared to the other study districts, but only in Jan-Feb 2023 and Jan-Feb 2024; otherwise, attrition was similar across the districts (Supplementary Table S8).

Among participants who attrited from at least one of the three main follow-ups, 43.4% (492/1134) rejoined the study at a later timepoint. The percentage of attriters from Jan-Feb 2023 and Oct 2023 who rejoined the study at the subsequent timepoint were 40.1% and 37% respectively as seen in the Supplementary Figure S2. Nearly 35% (65/186) of attrited households rejoined the study where both participants from the same household presented at the same rejoining timepoint. The average rate of rejoining across all villages was 43.3% (SD: 11.6%, range: 21.1%–68.2%). The percentage of participants who rejoined after an attrition event was lowest in Eastern Uganda (Pakwach 49.2%, 177/360; Buliisa 45.5%, 185/407; Mayuge 35.4%, 130/367).

### Determinants of overall attrition

Table 1 provides unadjusted associations of variables considered for the attrition analysis. Variables related to past IDs or any disease, religion, social status, preventative health behaviours, and spatial factors of district and distance to drug shops were not selected in univariable models.

Figure 2 shows adjusted results for the selected covariates for overall attrition. Individuals who belonged to households where other members of their household had an NCD were 28.7% more likely to miss at least one study timepoint when compared to individuals who were in households with no other members with NCDs (CI 1.072-1.545). With each one year increase in age, participants were 0.9% less likely to have an attrition event (CI 0.985-0.997). Higher levels of educational attainment were positively associated with attrition (7.7%; CI 1.047-1.108). Individuals whose household head owned their home were 22.7% less likely to have an attrition event when compared to individuals from rented homes (CI 0.599-0.998). Each one member increase in household size resulted in a 7.7% lower likelihood of having an attrition event (CI 0.863-0.987). Nearly all attrition clustering occurred within households (ICC 0.372) as opposed to within villages (ICC 0.22). When the model was rerun including the drug efficacy follow-up of March 2022 (Supplementary Table S1, Supplementary Figure S4), results remained robust. This timepoint therefore was excluded from further analysis.

**Fig. 2.**
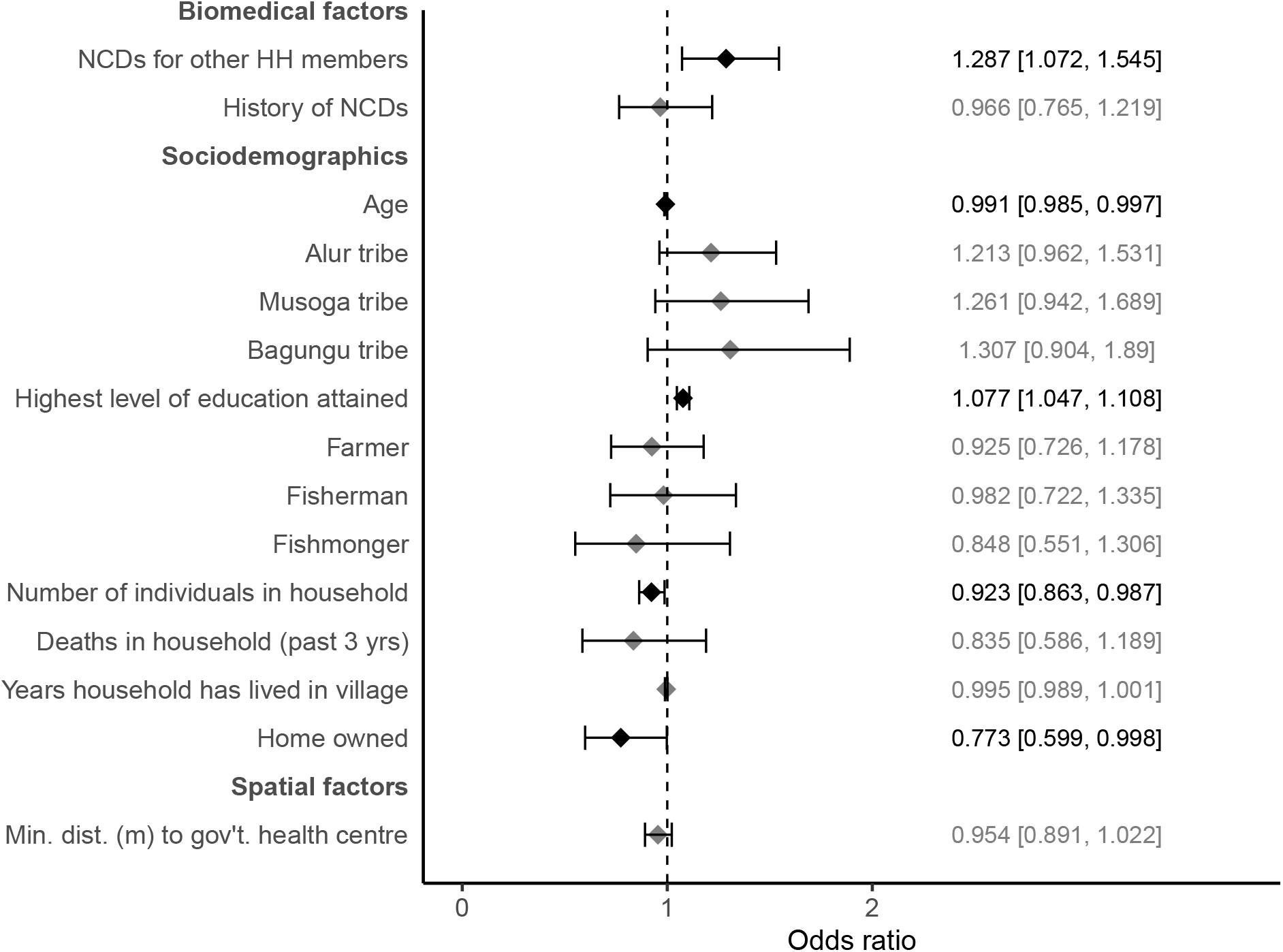
Overall attrition model. Logistic regression model for overall attrition of study participants (n=2844) with 95% confidence intervals calculated using household-level clustered standard errors (number of household clusters = 1445). VIFs *<* 10 for all variables. AUC for 10-fold cross-validation was 0.58. Results marked with a black diamond were significant and those marked in grey were non-significant.

Supplementary Figure S6 and Table S2 present the overall attrition at the household level (Figure S5). A higher level of education of the adult study participant was positively associated with household attrition (4.5%; CI 1.003-1.089). Households were 43.6% less likely to have an attrition event if there was someone in the household with social status as compared to households in which no members had social status (CI 0.332-0.915). For every 1 km closer a household was to a government health centre, the likelihood of missing a data collection timepoint decreased by 9.7% (CI 0.818-0.994).

### Seasonal variation in attrition and time to first attrition event

Supplementary Figures S7,S8, andS9 illustrate the relationship between participant characteristics and the seasonal variation of attrition with additional information provided in Supplementary Tables S3,S4, andS5. Higher education was associated with an increased likelihood of attrition, while a larger household size was linked to a reduced likelihood of attrition across all three major timepoints. Age was found to reduce the likelihood of attrition in Oct 2023 and Jan-Feb 2024, but it was not significantly associated with attrition in Jan-Feb 2023. Home ownership reduced the likelihood of attrition at the first major follow-up timepoint (Jan-Feb 2023), but not at other timepoints. In Oct 2023, the presence of NCDs among household members increased the likelihood of attrition, while proximity to a government health centre reduced it at a similar percentage (8.5%; CI 0.846–0.991) as the overall attrition model. Notably, this proximity variable was specific to the Oct 2023 timepoint and did not appear as a significant predictor in the overall attrition model at the individual level. The residuals of the logistic regression models for different time points were strongly correlated, with Pearson correlation coefficients of 0.42, 0.36, and 0.49 (all p <0.001) (Supplementary Figure S12).

The determinants of attrition became less distinguishable as the study matured. For the time to the first attrition event, there was a decline in the number of significant predictors at each timepoint as the study progressed (Supplementary Table S6, Supplementary Figure S10). In Jan-Feb 2023, many variables were significant for the first attrition event for an individual at that timepoint, including NCDs for other household members, sociodemographics of age, education, household size, home ownership, and the spatial variable of proximity to a government health centre. By Jan-Feb 2024, not a single observed variable was significant for the first event of attrition at that timepoint.

### Determinants of rejoining

Participants who attrited were not necessarily drop-outs from the study; individuals often returned to later study timepoints. The observed sequences of the attendance status of clinical participants at each timepoint along with frequencies of occurrence, as derived from study data, are shown in Figure 3. The most common sequence was attendance at all the timepoints, while the least common sequence was attendance during the baseline surveys in Jan-Feb 2022 and the most recent follow-up of Jan-Feb 2024 with non-attendance in between those years.

**Fig. 3.**
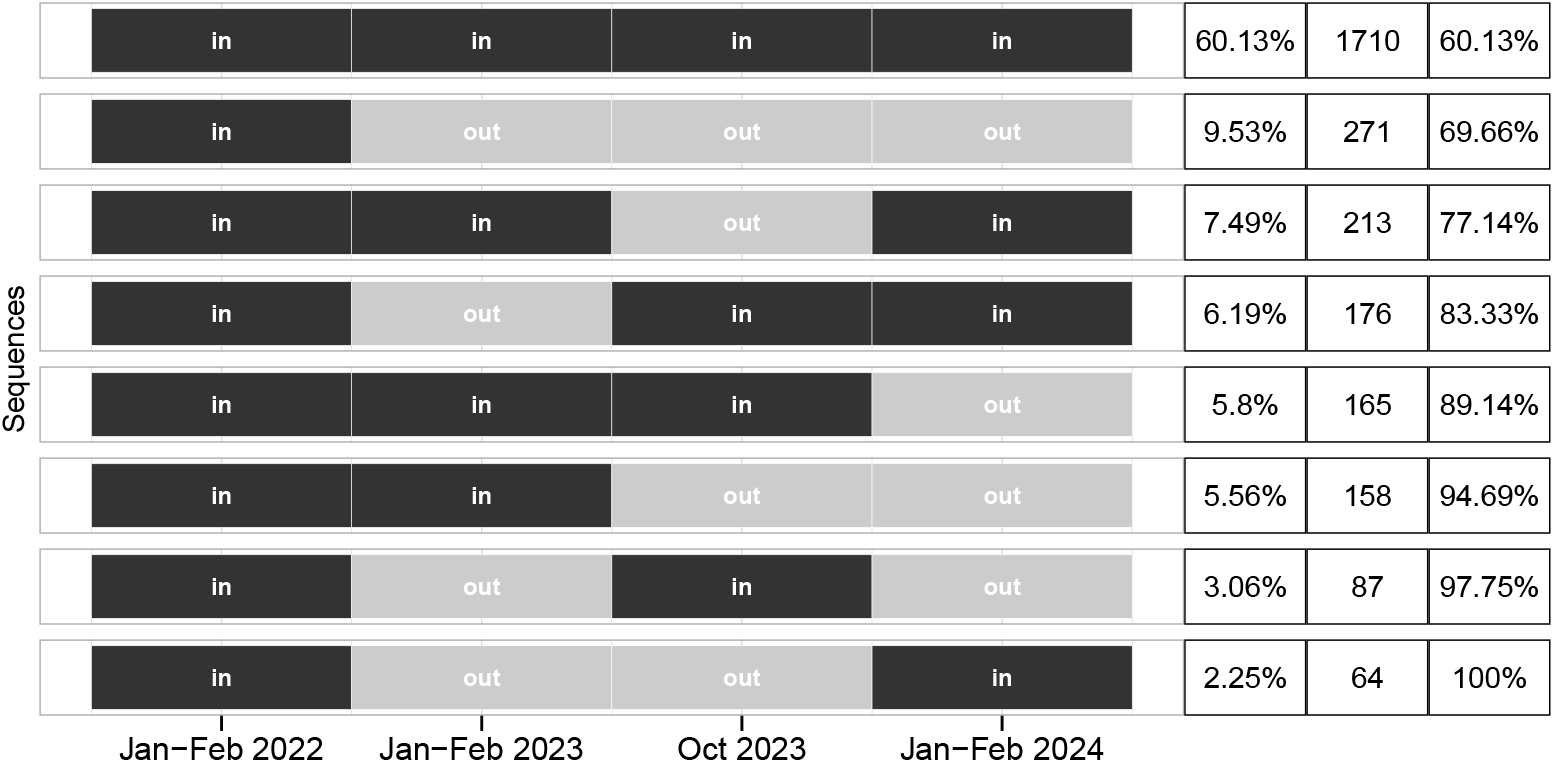
Attendance sequences. Sequences of participant attendance status during the main study timepoints.

There were few differences between rejoiners and individuals who stayed out of the study (Figure S11, Supplementary Table S7). Participants were 93.8% (CI 1.337-2.808) more likely to rejoin the study after having had an attrition event than to always stay out if they belonged to the Alur tribe, and 86.2% (CI 1.077-3.217) more likely if they belonged to the Bagungu tribe compared to participants belonging to tribes other than the three major tribes. Participants from households that experienced a death in the three years prior to Jan-Feb 2022 were 52.2% (CI 0.241-0.949) less likely to rejoin the study compared to those from households with no deaths during that period. While only these two variables were significant in distinguishing between rejoiners and those who remained out, the overall attrition model revealed starker differences between attrited and non-attrited participants, encompassing biomedical characteristics, sociodemographics, and spatial variables.

### Participant knowledge and later attrition

Information on exposures and outcomes provided to participants at each timepoint did not influence study attendance at a later timepoint (Supplementary Table S9). Concerning attrition bias, no associations of study outcomes with attrition were found. For study exposures, no consistent associations of infection across timepoints with attrition were found. Only *S. mansoni* infection from Jan-Feb 2023 was unreliably associated with attrition in Jan-Feb 2024 as infection status was borderline significant (p-value=0.047) and infection intensity was insignificant (p-value > 0.05).

## Discussion

Participant attrition in community-based cohort studies in sub-Saharan Africa has not been extensively explored outside of specialist populations, such as those for PLHIV or TB studies. Our study examines attrition from a general population cohort. We investigated 2844 individuals aged 5-92 within SchistoTrack.The study population represented diverse disease histories including healthy individuals, a wide range of socioeconomic statuses, differing levels of access to health care, distinct geographies, and varying levels of human mobility. Here we show the complexity of attrition metrics and provide a comprehensive framework for how future cohort studies can understand and monitor attrition. The detailed set of attrition rates provided may guide the choice of sample sizes needed for the design of future community-based cohorts in rural sub-Saharan Africa. Attrition was relatively stable ranging from 21-24.8% over three years and the profiles of individuals likely to have an attrition event were ascertained within only one year of the study.

Community engagement and tailored incentives may influence attrition by off-setting opportunity costs that participants may encounter when foregoing work or other activities to participate in a study ^19^. In our study, community engagement included not only the communication of medical findings, but also the provision of medical referrals and transport for urgent care. This approach resulted in almost no refusals for study participation for individuals across the three years. For incentives, there was a shift in SchistoTrack from 10,000 UGX to large household-size bars of soap.

The shift from in-cash to in-kind incentives, based on community feedback, did not influence participant attrition. This finding is further supported by the stable attrition rates observed across the timepoints. Participants were made aware of the incentive before each timepoint during sensitisation activities and participant mobilisation. Our finding suggests that our incentives were adequate and the type of incentive may be irrelevant in the context of broader study factors. This latter point has been shown in HIV cohorts ^20^ where low attrition rates were maintained by focusing on community interests in meetings and activities, along with adjustments to study team working hours. The provision of standard medical care and transportation reimbursement also has been shown to reduce attrition ^10^ and is essential for addressing participant needs ^21^. Hence, there is a complex interplay of community engagement and incentives where the former is likely to be more important than the latter for successful participant retention.

The provision of individual medical results from research studies within large-scale cohorts or Biobanks is uncommon ^22^. Yet, in extremely poor settings such as the context of SchistoTrack where care can be limited and the study may be the key source of healthcare engagement for the individual, it is an ethical imperative to share such findings with the study participants and local health workers ^21^. We found no evidence that sharing such findings will deter (or potentially encourage if the individual believes they are to receive some special care, conveying a therapeutic misconception ^23^) from participating in the study. Our experience within SchistoTrack demonstrates that understanding the study population, engaging them in meaningful dialogue, and responding to their needs is essential for supporting study participation and maintaining rigorous cohort designs.

The concept of attrition is inherently complex, especially in populations where individuals may rejoin the study after dropping out. Traditional views of attrition as a definitive dropout event do not fully capture the dynamics within highly mobile communities. Key factors associated with attrition, such as household size and home ownership are indicative of stability and settlement, while variables like age and education might be linked to possible migrations as individuals leave rural areas in search of opportunities in urban centres ^24^. The importance of household stability aligns with findings from an observational study for vaccine trials ^12^ where length of stay in the community was a significant factor for retention. However, in our study, these factors were only borderline significant, underscoring the need to carefully consider the minimal residency requirements when using participant or household stability as a study inclusion criterion. Households recruited within SchistoTrack are only required to have individuals who spend at least six months of the year within their village.

In our study, we observed that the profile of attriters could be largely established during the first follow-up, as the number of significant factors associated with a first attrition event decreased over time. Initially, the heterogeneity captured by our variables likely explained much of the observed attrition, but as the study progressed, the remaining instances may have been driven by random or unobserved characteristics, making it harder to pinpoint clear predictors. Alternatively, one possible limitation was simply there were fewer participants left to attrite (and predict) at later timepoints. A key consideration is whether our static treatment of baseline variables overlooked the impact of time-varying covariates. For example, while variables such as the presence of NCDs are likely stable over time, other variables such as household composition, occupation, and educational attainment, could evolve and influence attrition in ways not captured by our analysis. Future research could explore time-varying covariate modelling ^25^.

Our study challenges several assumptions about hard-to-reach populations, seasonal attrition, and the concept of true dropout in longitudinal, community-based studies. Although fishing communities are often considered difficult to track due to their mobility ^26 27^, our cohort included fishing populations and being a fisherman was never a discriminatory factor for predicting an attrition event either overall, by season, by household, or the time to the first attrition event. This finding suggests that populations typically perceived as challenging to engage may not require the stringent exclusion criteria, profiling for added community engagement or incentives, or oversampling in study designs to mitigate attrition. The lack of need for such adjustments is especially important considering that over 40% of attriters will rejoin. Despite high cross-border movements with the Democratic Republic of Congo ^28^, attrition rates were similar across Eastern and Western Uganda.

Attrition within community-based cohorts in sub-Saharan Africa is complex yet manageable with effective community engagement and metrics suited for understanding the determinants of attrition. We provide a starting set of statistics for other researchers to design recruitment and sampling strategies for community-based cohorts as well as analytical and conceptual frameworks for evaluating attrition in a manner that assesses effects on study outcomes and prioritises participant needs. If replicated in other studies, the attrition findings and retention strategies from SchistoTrack could help to construct guidelines for the robust design of large-scale community-based cohorts in rural sub-Saharan Africa.

## Data Availability

Participant data is not available due to the identifiable nature of the participant characteristics and ongoing nature of the cohort. Code is available from the authors upon request.

## Declarations

### Ethics approval

Data collection and use were reviewed and approved by Oxford Tropical Research Ethics Committee (OxTREC 509-21), Vector Control Division Research Ethics Committee of the Uganda Ministry of Health (VCDREC146), and Uganda National Council of Science and Technology (UNCST HS 1664ES).

## Acknowledgements

We thank the SchistoTrack Group for their valuable feedback during group meetings and informal exchanges, especially Fabian Reitzug and Lauren Wilburn for their insightful contributions. Special thanks to Saadiyah Mayet for her foundational work on the construction of some of the variables used in this study. We also thank the Ugandan SchistoTrack teams, including surveyors, technicians, nurses, sonographers, malacologists, HIV counsellors, and auxiliary workers, for their dedicated efforts. A special acknowledgment is due to the study participants, village health team members, local government nurses and surveyors, district leadership, and the Ministry of Health leadership, particularly at the Division of Vector Borne Diseases and Neglected Tropical Diseases. The successful execution of this research was made possible through their collective commitment and support.

## Author contributions

Conceptualization: GFC. Data curation: CP, BT, BN, NBK, GFC. Formal analysis: CP. Funding acquisition: GFC. Investigation: CP and GFC. Methodology: CP and GFC. Project administration: NBK. and GFC. Resources: NBK. and GFC. Software: GFC. Supervision: GFC. Validation: CP. Visualization: CP. Writing – original draft: CP. Writing – review & editing: CP, BT, BN, NBK, and GFC.

## References

1. Shadish WR, Luellen JK. Attrition. Wiley StatsRef: Statistics Reference Online. 2014.

2. Nunan D, Aronson J, Bankhead C. Catalogue of bias: attrition bias. BMJ evidence-based medicine. 2018;23(1):21–2.

3. Outes-Leon I, Dercon S. Survey attrition and attrition bias in Young Lives. Updated Publisher’s version, Young Lives Technical Note. 2009.

4. Akondeng C, Njamnshi WY, Mandi HE, et al. Community engagement in research in sub-Saharan Africa: approaches, barriers, facilitators, ethical considerations and the role of gender–a systematic review protocol. BMJ open. 2022;12(5):e057922.

5. Mousavian G, Ghalekhani N, Tavakoli F, et al. Proportion and reasons for loss to follow-up in a cohort study of people who inject drugs to measure HIV and HCV incidence in Kerman, Iran. Substance Abuse Treatment, Prevention, and Policy. 2021;16:1–8.

6. Fernández LG, Firima E, Robinson E, et al. Community-based care models for arterial hypertension management in non-pregnant adults in sub-Saharan Africa: a literature scoping review and framework for designing chronic services. BMC public health. 2022;22(1):1126.

7. Mengesha EW, Tesfaye TD, Boltena MT, et al. Effectiveness of community-based interventions for prevention and control of hypertension in sub-Saharan Africa: A systematic review. PLOS Global Public Health. 2024;4(7).

8. Kibuule D, Aiases P, Ruswa N, et al. Predictors of loss to follow-up of tuberculosis cases under the DOTS programme in Namibia. ERJ open research. 2020;6(1).

9. Frijters EM, Hermans LE, Wensing AM, et al. Risk factors for loss to follow-up from antiretroviral therapy programmes in low-income and middle-income countries. Aids. 2020;34(9):1261–88.

10. Kamya MR, Arinaitwe E, Wanzira H, et al. Malaria transmission, infection, and disease at three sites with varied transmission intensity in Uganda: implications for malaria control. The American journal of tropical medicine and hygiene. 2015;92(5):903.

11. Andolina C, Rek JC, Briggs J, et al. Sources of persistent malaria transmission in a setting with effective malaria control in eastern Uganda: a longitudinal, observational cohort study. The Lancet Infectious Diseases. 2021;21(11):1568–78.

12. Kiwanuka N, Mpendo J, Nalutaaya A, et al. An assessment of fishing communities around Lake Victoria, Uganda, as potential populations for future HIV vaccine efficacy studies: an observational cohort study. BMC public health. 2014;14:1–9.

13. Nuffield Department of Population Health UoO. SchistoTrack: a prospective multimorbidity cohort;. [Accessed 6 Aug 2024]. Available: https://www.bdi.ox.ac.uk/research/schistotrack.

14. Carstensen B, Plummer M, Laara E, et al. Epi: A Package for Statistical Analysis in Epidemiology; 2024. R package version 2.53. Available: https://CRAN.R-project.org/package=Epi.

15. Williams RL. A note on robust variance estimation for cluster-correlated data. Biometrics. 2000;56(2):645–6.

16. Chatterjee S, Hadi AS. Regression analysis by example. John Wiley & Sons; 2013.

17. Arlot S, Celisse A. A survey of cross-validation procedures for model selection. Statistics Surveys. 2010.

18. Anjorin S, Nabatte B, Mpooya S, et al. The epidemiology of periportal fibrosis and relevance of current Schistosoma mansoni infection: a population-based, cross-sectional study. medRxiv. 2023:2023-09.

19. Wang H, Kenkel D, Graham ML, et al. Cost-effectiveness of a community-based cardiovascular disease prevention intervention in medically underserved rural areas. BMC Health Services Research. 2019;19:1–13.

20. Seeley J, Nakiyingi-Miiro J, Kamali A, et al. High HIV incidence and socio-behavioral risk patterns in fishing communities on the shores of Lake Victoria, Uganda. Sexually transmitted diseases. 2012:433–9.

21. Frischer SR, Ockenden ES, Reitzug F, et al. Conceptualising care pathways for neglected tropical diseases in sub-Saharan Africa: A systematic scoping review. medRxiv. 2024:2024-09.

22. Chen Z, Chen J, Collins R, et al. China Kadoorie Biobank of 0.5 million people: survey methods, baseline characteristics and long-term follow-up. International journal of epidemiology. 2011;40(6):1652–66.

23. Appelbaum PS, Roth LH, Lidz CW, et al. False hopes and best data: consent to research and the therapeutic misconception. In: Research Ethics. Routledge; 2017. p. 167–71.

24. Ginsburg C, Bocquier P, Béguy D, et al. Human capital on the move: Education as a determinant of internal migration in selected INDEPTH surveillance populations in Africa. Demographic Research. 2016;34:845.

25. Zhang Z, Reinikainen J, Adeleke KA, et al. Time-varying covariates and coefficients in Cox regression models. Annals of translational medicine. 2018;6(7).

26. Parker M, Allen T, Pearson G, et al. Border parasites: schistosomiasis control among Uganda’s fisherfolk. Journal of Eastern African Studies. 2012;6(1):98–123.

27. Bogart LM, Naigino R, Maistrellis E, et al. Barriers to linkage to HIV care in Ugandan fisherfolk communities: a qualitative analysis. AIDS and Behavior. 2016;20:2464–76.

28. Bedford J, Akello G, Gercama I, et al. Uganda-DRC cross-border dynamics. F1000Research. 2020;9(390):390.

