## Supplementary material for "Maintaining community-based cohorts in sub-Saharan Africa: An evaluation of participant attrition in SchistoTrack"

### Supplementary information

#### S1 Supplementary methods

##### S1.1 Covariate definitions

Age was recorded as a continuous variable in years at the time of recruitment. Sex was a binary variable coded as male or female with male being the reference category. Tribe was categorised for the three majority tribes of Alur, Musoga, and Bagungu, with the reference category being all other tribes. Majority religion was a binary variable indicating whether the participant belonged to the predominant religion in their village, with not belonging as the reference category. The highest level of education attained was an ordinal variable ranging from 0 to 14, with 0 corresponding to no education; 1-7 to primary level education; 8-12 to senior level education; 13 to diploma, certificate, or some university education; and 14 to completed university education [1]. The reference level was 0, indicating no level of primary education was completed. Occupation was categorised into farmer, fisherman, and fishmonger with no occupations and other occupations as one reference category. The occupation variable was curated to capture the behaviour of populations at low and high risks of attrition based on assumed differences in mobility given fishermen move to areas with the best catches and farmers have designated set land for which to use for subsistence.

The home quality score variable was a continuous variable generated by ranking the materials of the roof, walls, and floor for each household on a scale from 1 to 4, and then summing these ranks. We followed Chami et al. [1] for the ranking of materials, with the order from lowest to highest quality as follows: for the roof, grass or papyrus, sticks, plastic, and metal; for the walls, mud and sticks, plastic, metal, and bricks or cement; and for the floor, mud, plastic, wood planks, and brick or cement. Household social status was a binary variable indicating whether one or more members of the participant's household members held a position in the local administration, with no members holding an administrative position as the reference category. The number of individuals in the household, indicating household size, was recorded as a count. Deaths in the household in the three years before recruitment were coded as a binary variable, with the reference category indicating the absence of deaths in the household. The number of years the household has lived in the village, suggestive of household stability, was recorded as a continuous variable. Home ownership was a binary variable, with the reference level being that the household head did not own their home and rented the home. The number of rooms in the home was recorded as a count, reflecting household density and living conditions as well as possible wealth.

Household-level variables related to water, sanitation, and hygiene (WASH) were considered in the analysis as preventive health measures potentially influencing participation in the study. An improved drinking water source [2] was included as a binary variable, with sources such as a protected well or spring, borehole, village tap, or rainwater classified as improved, while other sources served as the reference category. Household water treatment [3] was included as a binary variable, with no water treatment or efforts to purify or make the water safe for consumption after collection as the reference value. Methods of water treatment considered included boiling, adding bleach or chlorine, straining through a cloth, using a water filter, solar disinfection, washing the jerrycan used for storing water with soap, and letting the water stand and settle. Having an improved sanitation facility [2], such as a flush toilet or a covered latrine with privacy, was another binary indicator, with the absence of such a facility as the reference level. Additionally, having provisions for washing hands with water and soap was included as a binary variable, with the lack of such provisions as the reference.

For individual-level biomedical factors, the presence of any symptoms of physical or mental illness reported by the study participant within the one month before recruitment was coded as a binary variable, with the reference level being the absence of any such symptoms. The history of IDs was coded as a binary variable indicating whether the participant had ever been told by a doctor or health worker that they had one of the following IDs of bilharzia, brucellosis, cholera, complicated/severe malaria, COVID-19, diarrhea, dysentery, encephalitis, gut worms, hemorrhagic fever, hepatitis B, hepatitis C, HIV/AIDS, leprosy, measles, meningitis, pertussis, pneumonia, scabies, sepsis, TB, tetanus, trachoma, and uncomplicated malaria. Having any sexually transmitted infection, such as syphilis, gonorrhea, chlamydia, genital herpes, human papillomavirus, HIV, or other infections, was also considered a condition for participants aged 20 or above. NCDs were assessed using modified versions of the World Health Organization (WHO) STEPwise approach to NCD risk factor surveillance (STEPS) [4]. The history of NCDs was a binary variable recorded only for participants aged 20 or above who had ever been told by a doctor or health worker that they had one of the following conditions of anxiety, asthma, cancer, depression, eczema, psychosis, sickle cell disease, heart disease, raised blood pressure, raised cholesterol, or raised blood sugar, or if they were a current smoker or drinker. The reference category for the history of NCDs was having never had or currently having none of the above conditions to the knowledge of the participant. The history of any disease for the participant was a binary indicator based on the previously defined variables of history of IDs and history of NCDs, with the reference value being never having had any of the health conditions.

The spatial factors of distances to drug shops and health centres were calculated using the `raster` package in R [5], as the shortest distance between the waypoint coordinates of the drug shops and health centres, collected by surveyors in each district, and the waypoint coordinates of each household in the respective district. Households were iteratively compared to each drug shop and health center within their district to determine the closest distances. The positional accuracy of the locations was constrained by the average waypoint accuracy of 5 metres for the drug shops and health centres, and 10 metres for the household locations, as recorded in the ODK surveys. These spatial factors were considered to account for variations in access to health services and infrastructure, which could influence participation in the study.

For the analysis of household-level attrition, the inclusion of variables at this level aimed to capture the broader socio-economic and demographic context that might influence the likelihood of households dropping out of the study. By considering variables that reflect the overall characteristics of the household, our analysis sought to identify patterns and factors that could impact household stability and participation. In addition to all the household-level variables described earlier, the analysis also incorporated the individual-level variables of the highest level of education attained and occupation. However, for these variables, data were drawn specifically from the adult study participant within the household. The highest level of education attained was coded as an ordinal variable for the adult study participant exactly as described at the individual level. Occupation was coded as previously described at the individual level to capture the primary economic activity of the adult participant, which could reflect the household's economic stability and risk of attrition. This approach ensured that the analysis accounted for both the collective attributes of the household and the individual characteristics of its adult study participant, offering a comprehensive view of the factors influencing household-level attrition.

##### S1.2 Variable definitions

PPF was diagnosed with point-of-care ultrasound following the Niamey Protocol [6] as described in detail elsewhere [7]. All detected patterns of PPF were documented, and the most severe pattern observed for each participant was used to create a binary outcome variable. In this classification, any participant with a pattern in the C-F range was categorised as having PPF, while patterns in the A-B range were classified as no PPF. Schistosomiasis-related referrals were mainly due to low Hb counts, severe liver fibrotic disease from ultrasound scans, hepatitis-like livers from ultrasound scans, nurse-determined urgent needs for blood transfusion, self-reported history of vomiting blood, portal hypertension, splenomegaly, hepatomegaly, liver cirrhosis, need for PZQ treatment, ascites, and observed liver masses from ultrasound as unclear whether schistosomiasis-related or due to other causes. Schistosome infection status was ascertained using Kato-Katz (KK) microscopy [8] in which two thick smear slides were prepared from a single stool sample, and the two slides were examined by two different technicians. The results from the slides were then averaged and multiplied by 24 to estimate the number of eggs per gram (EPG) of faeces. A positive KK infection status was determined when EPG>=1. For infection intensity, WHO categories [9] were applied, where the classifications were no infection (0 EPG), low (1-99 EPG), moderate (100-399 EPG), and heavy (400+ EPG).

##### S1.3 Additional statistical models

Descriptive statistics were used to compare participant and household characteristics between attrited and non-attrited groups using chi-squared tests for categorical variables and t-tests for numerical variables. Logistic regression models were used to investigate inclusion of the March 2022 timepoint, household attrition, seasonal variation in attrition, time of first attrition, and rejoining the study. The same model-building procedures outlined in the main text were applied to all analyses. Except for the household-level model, variables selected in the overall attrition model were used in all participant-level models. Also, clustered standard errors at the household level were used to account for the paired sampling design in all individual-level models except the time of first attrition multinomial model.

To investigate whether inclusion of the short-term March 2022 timepoint made a significant difference, logistic regression for attrited and non-attrited participants on variables selected for overall attrition including the short-term follow-up timepoint after baseline was performed. For seasonal variation in attrition, logistic regression models were applied separately at each of the three main follow-up timepoints (Jan-Feb 2023, Oct 2023, and Jan-Feb 2024) using the same predictors as in the overall attrition model. To explore first attrition timing, multinomial logistic regression was employed, comparing participants who always stayed in the study to those who first attrited at one of the three timepoints. For rejoining the study, logistic regression was used to compare participants who had had an attrition event but later rejoined and those who remained out of the study.

#### S2 Supplementary tables

**Table S1.** Summary statistics of potential predictors of attrition, including the March 2022 drug efficacy timepoint.

| | Total | Attrited | Non-attrited |
|---|---|---|---|
| | (N=2844) | (N=1175) | (N=1669) |
| BIOMEDICAL FACTORS | | | |
| Any symptoms | 129 (4.5%) | 45 (3.8%) | 84 (5.0%) |
| IDs for other HH members | 1739 (61.1%) | 708 (60.3%) | 1031 (61.8%) |
| NCDs for other HH members | 965 (33.9%) | 431 (36.7%) | 534 (32.0%) |
| History of IDs | 1405 (49.4%) | 566 (48.2%) | 839 (50.3%) |
| History of NCDs | 543 (19.1%) | 203 (17.3%) | 340 (20.4%) |
| History of any disease | 1584 (55.7%) | 642 (54.6%) | 942 (56.4%) |
| SOCIODEMOGAPHICS | | | |
| Age (Mean (SD)) | 24.6 (18.0) | 22.9 (16.5) | 25.8 (18.8) |
| Sex - Female | 1565 (55.0%) | 655 (55.7%) | 910 (54.5%) |
| Tribe | | | |
| Alur | 1550 (54.5%) | 647 (55.1%) | 903 (54.1%) |
| Bagungu | 235 (8.3%) | 115 (9.8%) | 120 (7.2%) |
| Musoga | 461 (16.2%) | 190 (16.2%) | 271 (16.2%) |
| Other | 598 (21.0%) | 223 (19.0%) | 375 (22.5%) |
| Majority religion | 2049 (72.0%) | 838 (71.3%) | 1211 (72.6%) |
| Highest level of education attained (Mean (SD)) | 3.45 (2.83) | 3.72 (2.88) | 3.27 (2.78) |
| Occupation | | | |
| Farmer | 508 (17.9%) | 178 (15.1%) | 330 (19.8%) |
| Fisherman | 243 (8.5%) | 106 (9.0%) | 137 (8.2%) |
| Fishmonger | 108 (3.8%) | 41 (3.5%) | 67 (4.0%) |
| None/Other | 1985 (69.8%) | 850 (72.3%) | 1135 (68.0%) |
| Home quality score (Mean (SD)) | 6.38 (3.56) | 6.49 (3.64) | 6.30 (3.49) |
| Household social status | 285 (10.0%) | 106 (9.0%) | 179 (10.7%) |
| Number of individuals in household (Mean (SD)) | 3.63 (1.38) | 3.55 (1.26) | 3.69 (1.46) |
| Deaths in household (past 3 yrs) | 184 (6.5%) | 64 (5.4%) | 120 (7.2%) |
| Years household has lived in village (Mean (SD)) | 18.9 (14.9) | 17.7 (14.4) | 19.8 (15.2) |
| Home owned | 2403 (84.5%) | 967 (82.3%) | 1436 (86.0%) |
| Number of rooms (Mean (SD)) | 2.17 (1.23) | 2.16 (1.27) | 2.18 (1.20) |
| WATER, SANITATION AND HYGIENE (WASH) | | | |
| Improved drinking water source | 1667 (58.6%) | 682 (58.0%) | 985 (59.0%) |
| Household treats drinking water | 643 (22.6%) | 259 (22.0%) | 384 (23.0%) |
| Improved sanitation facility in home | 1742 (61.3%) | 695 (59.1%) | 1047 (62.7%) |
| Basic hygiene facility in home | 311 (10.9%) | 117 (10.0%) | 194 (11.6%) |
| SPATIAL FACTORS | | | |
| Min. dist. (km) to drug shop (Mean (SD)) | 0.937 (1.15) | 0.893 (1.15) | 0.968 (1.15) |
| Min. dist. (km) to gov't. health centre (Mean (SD)) | 2.30 (1.33) | 2.23 (1.28) | 2.34 (1.36) |
| District | | | |
| Buliisa | 947 (33.3%) | 419 (35.7%) | 528 (31.6%) |
| Mayuge | 953 (33.5%) | 380 (32.3%) | 573 (34.3%) |
| Pakwach | 944 (33.2%) | 376 (32.0%) | 568 (34.0%) |

**Table S2.** Summary statistics of potential predictors of HH attrition.

| | Total | HH attrited | HH non-attrited | P-value |
|---|---|---|---|---|
| | (N=1399) | (N=316) | (N=1083) | ($\chi^2$/t-test) |
| SOCIODEMOGAPHICS | | | | |
| Highest level of education attained (adult study participant) | | | | 0.0222 |
| Mean (SD) | 4.42 (3.17) | 4.78 (3.22) | 4.31 (3.15) | |
| Occupation (adult study participant) | | | | 0.129 |
| Farmer | 481 (34.4%) | 93 (29.4%) | 388 (35.8%) | |
| Fisherman | 236 (16.9%) | 52 (16.5%) | 184 (17.0%) | |
| Fishmonger | 107 (7.6%) | 29 (9.2%) | 78 (7.2%) | |
| None/Other | 575 (41.1%) | 142 (44.9%) | 433 (40.0%) | |
| Home quality score | | | | 0.773 |
| Mean (SD) | 6.40 (3.56) | 6.46 (3.64) | 6.39 (3.54) | |
| Household social status | 142 (10.2%) | 20 (6.3%) | 122 (11.3%) | 0.0143 |
| Number of individuals in household | | | | 0.0618 |
| Mean (SD) | 3.64 (1.38) | 3.53 (1.20) | 3.68 (1.43) | |
| Deaths in household (past 3 yrs) | 90 (6.4%) | 12 (3.8%) | 78 (7.2%) | 0.0413 |
| Years household has lived in village | | | | <0.001 |
| Mean (SD) | 18.9 (14.9) | 16.3 (13.9) | 19.7 (15.1) | |
| Home owned | 1181 (84.4%) | 249 (78.8%) | 932 (86.1%) | 0.00235 |
| Number of rooms | | | | 0.14 |
| Mean (SD) | 2.18 (1.24) | 2.09 (1.24) | 2.21 (1.23) | |
| WATER, SANITATION AND HYGIENE (WASH) | | | | |
| Improved drinking water source | 822 (58.8%) | 186 (58.9%) | 636 (58.7%) | 1 |
| Household treats drinking water | 316 (22.6%) | 67 (21.2%) | 249 (23.0%) | 0.553 |
| Improved sanitation facility in home | 857 (61.3%) | 180 (57.0%) | 677 (62.5%) | 0.0861 |
| Basic hygiene facility in home | 154 (11.0%) | 28 (8.9%) | 126 (11.6%) | 0.199 |
| SPATIAL FACTORS | | | | |
| Min. dist. (km) to drug shop | | | | 0.51 |
| Mean (SD) | 0.934 (1.15) | 0.895 (1.19) | 0.945 (1.14) | |
| Min. dist. (km) to gov't. health centre | | | | 0.0324 |
| Mean (SD) | 2.30 (1.33) | 2.17 (1.26) | 2.34 (1.35) | |
| District | | | | 0.00657 |
| Buliisa | 464 (33.2%) | 128 (40.5%) | 336 (31.0%) | |
| Pakwach | 471 (33.7%) | 97 (30.7%) | 374 (34.5%) | |
| Mayuge | 464 (33.2%) | 91 (28.8%) | 373 (34.4%) | |

**Table S3.** Summary statistics of potential predictors for attrition in Jan-Feb 2023.

| | Total | Non-attrited | Attrited |
|---|---|---|---|
| | (N=2844) | (N=2246) | (N=598) |
| BIOMEDICAL FACTORS | | | |
| Any symptoms | 129 (4.5%) | 102 (4.5%) | 27 (4.5%) |
| IDs for other HH members | 1739 (61.1%) | 1373 (61.1%) | 366 (61.2%) |
| NCDs for other HH members | 965 (33.9%) | 748 (33.3%) | 217 (36.3%) |
| History of IDs | 1405 (49.4%) | 1116 (49.7%) | 289 (48.3%) |
| History of NCDs | 543 (19.1%) | 434 (19.3%) | 109 (18.2%) |
| History of any disease | 1584 (55.7%) | 1259 (56.1%) | 325 (54.3%) |
| SOCIODEMOGAPHICS | | | |
| Age (Mean (SD)) | 24.6 (18.0) | 25.1 (18.3) | 22.7 (16.4) |
| Sex - Female | 1565 (55.0%) | 1224 (54.5%) | 341 (57.0%) |
| Tribe | | | |
| Alur | 1550 (54.5%) | 1216 (54.1%) | 334 (55.9%) |
| Bagungu | 235 (8.3%) | 172 (7.7%) | 63 (10.5%) |
| Musoga | 461 (16.2%) | 364 (16.2%) | 97 (16.2%) |
| Other | 598 (21.0%) | 494 (22.0%) | 104 (17.4%) |
| Majority religion | 2049 (72.0%) | 1625 (72.4%) | 424 (70.9%) |
| Highest level of education attained (Mean (SD)) | 3.45 (2.83) | 3.36 (2.78) | 3.81 (2.98) |
| Occupation | | | |
| Farmer | 508 (17.9%) | 430 (19.1%) | 78 (13.0%) |
| Fisherman | 243 (8.5%) | 184 (8.2%) | 59 (9.9%) |
| Fishmonger | 108 (3.8%) | 84 (3.7%) | 24 (4.0%) |
| None/Other | 1985 (69.8%) | 1548 (68.9%) | 437 (73.1%) |
| Home quality score (Mean (SD)) | 6.38 (3.56) | 6.38 (3.56) | 6.39 (3.56) |
| Household social status | 285 (10.0%) | 237 (10.6%) | 48 (8.0%) |
| Number of individuals in household (Mean (SD)) | 3.63 (1.38) | 3.69 (1.43) | 3.41 (1.15) |
| Deaths in household (past 3 yrs) | 184 (6.5%) | 159 (7.1%) | 25 (4.2%) |
| Years household has lived in village (Mean (SD)) | 18.9 (14.9) | 19.5 (15.1) | 17.0 (14.2) |
| Home owned | 2403 (84.5%) | 1930 (85.9%) | 473 (79.1%) |
| Number of rooms (Mean (SD)) | 2.17 (1.23) | 2.21 (1.26) | 2.04 (1.11) |
| WATER, SANITATION AND HYGIENE (WASH) | | | |
| Improved drinking water source | 1667 (58.6%) | 1306 (58.1%) | 361 (60.4%) |
| Household treats drinking water | 643 (22.6%) | 515 (22.9%) | 128 (21.4%) |
| Improved sanitation facility in home | 1742 (61.3%) | 1413 (62.9%) | 329 (55.0%) |
| Basic hygiene facility in home | 311 (10.9%) | 266 (11.8%) | 45 (7.5%) |
| SPATIAL FACTORS | | | |
| Min. dist. (km) to drug shop (Mean (SD)) | 0.937 (1.15) | 0.942 (1.14) | 0.919 (1.20) |
| Min. dist. (km) to gov't. health centre (Mean (SD)) | 2.30 (1.33) | 2.33 (1.35) | 2.18 (1.22) |
| District | | | |
| Buliisa | 947 (33.3%) | 711 (31.7%) | 236 (39.5%) |
| Mayuge | 953 (33.5%) | 769 (34.2%) | 184 (30.8%) |
| Pakwach | 944 (33.2%) | 766 (34.1%) | 178 (29.8%) |

**Table S4.** Summary statistics of potential predictors for attrition in Oct 2023.

| | Total | Non-attrited | Attrited |
|---|---|---|---|
| | (N=2844) | (N=2163) | (N=681) |
| BIOMEDICAL FACTORS | | | |
| Any symptoms | 129 (4.5%) | 102 (4.7%) | 27 (4.0%) |
| IDs for other HH members | 1739 (61.1%) | 1333 (61.6%) | 406 (59.6%) |
| NCDs for other HH members | 965 (33.9%) | 697 (32.2%) | 268 (39.4%) |
| History of IDs | 1405 (49.4%) | 1092 (50.5%) | 313 (46.0%) |
| History of NCDs | 543 (19.1%) | 443 (20.5%) | 100 (14.7%) |
| History of any disease | 1584 (55.7%) | 1234 (57.1%) | 350 (51.4%) |
| SOCIODEMOGAPHICS | | | |
| Age (Mean (SD)) | 24.6 (18.0) | 25.6 (18.6) | 21.2 (15.5) |
| Sex - Female | 1565 (55.0%) | 1179 (54.5%) | 386 (56.7%) |
| Tribe | | | |
| Alur | 1550 (54.5%) | 1162 (53.7%) | 388 (57.0%) |
| Bagungu | 235 (8.3%) | 171 (7.9%) | 64 (9.4%) |
| Musoga | 461 (16.2%) | 354 (16.4%) | 107 (15.7%) |
| Other | 598 (21.0%) | 476 (22.0%) | 122 (17.9%) |
| Majority religion | 2049 (72.0%) | 1556 (71.9%) | 493 (72.4%) |
| Highest level of education attained (Mean (SD)) | 3.45 (2.83) | 3.36 (2.81) | 3.75 (2.86) |
| Occupation | | | |
| Farmer | 508 (17.9%) | 416 (19.2%) | 92 (13.5%) |
| Fisherman | 243 (8.5%) | 190 (8.8%) | 53 (7.8%) |
| Fishmonger | 108 (3.8%) | 85 (3.9%) | 23 (3.4%) |
| None/Other | 1985 (69.8%) | 1472 (68.1%) | 513 (75.3%) |
| Home quality score (Mean (SD)) | 6.38 (3.56) | 6.38 (3.53) | 6.38 (3.64) |
| Household social status | 285 (10.0%) | 225 (10.4%) | 60 (8.8%) |
| Number of individuals in household (Mean (SD)) | 3.63 (1.38) | 3.68 (1.43) | 3.50 (1.21) |
| Deaths in household (past 3 yrs) | 184 (6.5%) | 148 (6.8%) | 36 (5.3%) |
| Years household has lived in village (Mean (SD)) | 18.9 (14.9) | 19.3 (14.8) | 17.8 (15.1) |
| Home owned | 2403 (84.5%) | 1848 (85.4%) | 555 (81.5%) |
| Number of rooms (Mean (SD)) | 2.17 (1.23) | 2.19 (1.20) | 2.12 (1.32) |
| WATER, SANITATION AND HYGIENE (WASH) | | | |
| Improved drinking water source | 1667 (58.6%) | 1276 (59.0%) | 391 (57.4%) |
| Household treats drinking water | 643 (22.6%) | 487 (22.5%) | 156 (22.9%) |
| Improved sanitation facility in home | 1742 (61.3%) | 1337 (61.8%) | 405 (59.5%) |
| Basic hygiene facility in home | 311 (10.9%) | 251 (11.6%) | 60 (8.8%) |
| SPATIAL FACTORS | | | |
| Min. dist. (km) to drug shop (Mean (SD)) | 0.937 (1.15) | 0.935 (1.13) | 0.944 (1.21) |
| Min. dist. (km) to gov't. health centre (Mean (SD)) | 2.30 (1.33) | 2.34 (1.34) | 2.17 (1.28) |
| District | | | |
| Buliisa | 947 (33.3%) | 698 (32.3%) | 249 (36.6%) |
| Mayuge | 953 (33.5%) | 743 (34.4%) | 210 (30.8%) |
| Pakwach | 944 (33.2%) | 722 (33.4%) | 222 (32.6%) |

**Table S5.** Summary statistics of potential predictors for attrition in Jan-Feb 2024.

| | Total | Non-attrited | Attrited |
|---|---|---|---|
| | (N=2844) | (N=2138) | (N=706) |
| BIOMEDICAL FACTORS | | | |
| Any symptoms | 129 (4.5%) | 103 (4.8%) | 26 (3.7%) |
| IDs for other HH members | 1739 (61.1%) | 1332 (62.3%) | 407 (57.6%) |
| NCDs for other HH members | 965 (33.9%) | 703 (32.9%) | 262 (37.1%) |
| History of IDs | 1405 (49.4%) | 1081 (50.6%) | 324 (45.9%) |
| History of NCDs | 543 (19.1%) | 436 (20.4%) | 107 (15.2%) |
| History of any disease | 1584 (55.7%) | 1214 (56.8%) | 370 (52.4%) |
| SOCIODEMOGAPHICS | | | |
| Age (Mean (SD)) | 24.6 (18.0) | 25.6 (18.6) | 21.4 (15.5) |
| Sex - Female | 1565 (55.0%) | 1176 (55.0%) | 389 (55.1%) |
| Tribe | | | |
| Alur | 1550 (54.5%) | 1194 (55.8%) | 356 (50.4%) |
| Bagungu | 235 (8.3%) | 171 (8.0%) | 64 (9.1%) |
| Musoga | 461 (16.2%) | 329 (15.4%) | 132 (18.7%) |
| Other | 598 (21.0%) | 444 (20.8%) | 154 (21.8%) |
| Majority religion | 2049 (72.0%) | 1552 (72.6%) | 497 (70.4%) |
| Highest level of education attained (Mean (SD)) | 3.45 (2.83) | 3.39 (2.83) | 3.66 (2.81) |
| Occupation | | | |
| Farmer | 508 (17.9%) | 410 (19.2%) | 98 (13.9%) |
| Fisherman | 243 (8.5%) | 193 (9.0%) | 50 (7.1%) |
| Fishmonger | 108 (3.8%) | 86 (4.0%) | 22 (3.1%) |
| None/Other | 1985 (69.8%) | 1449 (67.8%) | 536 (75.9%) |
| Home quality score (Mean (SD)) | 6.38 (3.56) | 6.30 (3.54) | 6.62 (3.60) |
| Household social status | 285 (10.0%) | 233 (10.9%) | 52 (7.4%) |
| Number of individuals in household (Mean (SD)) | 3.63 (1.38) | 3.68 (1.43) | 3.50 (1.23) |
| Deaths in household (past 3 yrs) | 184 (6.5%) | 143 (6.7%) | 41 (5.8%) |
| Years household has lived in village (Mean (SD)) | 18.9 (14.9) | 19.4 (15.0) | 17.6 (14.5) |
| Home owned | 2403 (84.5%) | 1828 (85.5%) | 575 (81.4%) |
| Number of rooms (Mean (SD)) | 2.17 (1.23) | 2.18 (1.24) | 2.14 (1.21) |
| WATER, SANITATION AND HYGIENE (WASH) | | | |
| Improved drinking water source | 1667 (58.6%) | 1265 (59.2%) | 402 (56.9%) |
| Household treats drinking water | 643 (22.6%) | 494 (23.1%) | 149 (21.1%) |
| Improved sanitation facility in home | 1742 (61.3%) | 1314 (61.5%) | 428 (60.6%) |
| Basic hygiene facility in home | 311 (10.9%) | 243 (11.4%) | 68 (9.6%) |
| SPATIAL FACTORS | | | |
| Min. dist. (km) to drug shop (Mean (SD)) | 0.937 (1.15) | 0.969 (1.16) | 0.840 (1.12) |
| Min. dist. (km) to gov't. health centre (Mean (SD)) | 2.30 (1.33) | 2.31 (1.35) | 2.25 (1.27) |
| District | | | |
| Buliisa | 947 (33.3%) | 698 (32.6%) | 249 (35.3%) |
| Mayuge | 953 (33.5%) | 690 (32.3%) | 263 (37.3%) |
| Pakwach | 944 (33.2%) | 750 (35.1%) | 194 (27.5%) |

**Table S6.** Summary statistics of potential predictors for first attrition.

| | Total | Always in | Jan-Feb 2023 | Oct 2023 | Jan-Feb 2024 |
|---|---|---|---|---|---|
| | (N=2844) | (N=1710) | (N=598) | (N=323) | (N=213) |
| BIOMEDICAL FACTORS | | | | | |
| Any symptoms | 129 (4.5%) | 85 (5.0%) | 27 (4.5%) | 10 (3.1%) | 7 (3.3%) |
| IDs for other HH members | 1739 (61.1%) | 1055 (61.7%) | 366 (61.2%) | 192 (59.4%) | 126 (59.2%) |
| NCDs for other HH members | 965 (33.9%) | 543 (31.8%) | 217 (36.3%) | 133 (41.2%) | 72 (33.8%) |
| History of IDs | 1405 (49.4%) | 860 (50.3%) | 289 (48.3%) | 151 (46.7%) | 105 (49.3%) |
| History of NCDs | 543 (19.1%) | 350 (20.5%) | 109 (18.2%) | 46 (14.2%) | 38 (17.8%) |
| History of any disease | 1584 (55.7%) | 967 (56.5%) | 325 (54.3%) | 170 (52.6%) | 122 (57.3%) |
| SOCIODEMOGAPHICS | | | | | |
| Age (Mean (SD)) | 24.6 (18.0) | 25.9 (18.9) | 22.7 (16.4) | 21.6 (15.7) | 24.0 (17.1) |
| Sex - Female | 1565 (55.0%) | 934 (54.6%) | 341 (57.0%) | 182 (56.3%) | 108 (50.7%) |
| Tribe | | | | | |
| Alur | 1550 (54.5%) | 920 (53.8%) | 334 (55.9%) | 190 (58.8%) | 106 (49.8%) |
| Bagungu | 235 (8.3%) | 127 (7.4%) | 63 (10.5%) | 27 (8.4%) | 18 (8.5%) |
| Musoga | 461 (16.2%) | 274 (16.0%) | 97 (16.2%) | 49 (15.2%) | 41 (19.2%) |
| Other | 598 (21.0%) | 389 (22.7%) | 104 (17.4%) | 57 (17.6%) | 48 (22.5%) |
| Majority religion | 2049 (72.0%) | 1241 (72.6%) | 424 (70.9%) | 239 (74.0%) | 145 (68.1%) |
| Highest level of education attained (Mean (SD)) | 3.45 (2.83) | 3.28 (2.79) | 3.81 (2.98) | 3.71 (2.72) | 3.50 (2.75) |
| Occupation | | | | | |
| Farmer | 508 (17.9%) | 336 (19.6%) | 78 (13.0%) | 48 (14.9%) | 46 (21.6%) |
| Fisherman | 243 (8.5%) | 145 (8.5%) | 59 (9.9%) | 23 (7.1%) | 16 (7.5%) |
| Fishmonger | 108 (3.8%) | 70 (4.1%) | 24 (4.0%) | 9 (2.8%) | 5 (2.3%) |
| None/Other | 1985 (69.8%) | 1159 (67.8%) | 437 (73.1%) | 243 (75.2%) | 146 (68.5%) |
| Home quality score (Mean (SD)) | 6.38 (3.56) | 6.31 (3.50) | 6.39 (3.56) | 6.33 (3.73) | 6.98 (3.68) |
| Household social status | 285 (10.0%) | 184 (10.8%) | 48 (8.0%) | 34 (10.5%) | 19 (8.9%) |
| Number of individuals in household (Mean (SD)) | 3.63 (1.38) | 3.70 (1.46) | 3.41 (1.15) | 3.64 (1.27) | 3.70 (1.41) |
| Deaths in household (past 3 yrs) | 184 (6.5%) | 124 (7.3%) | 25 (4.2%) | 15 (4.6%) | 20 (9.4%) |
| Years household has lived in village (Mean (SD)) | 18.9 (14.9) | 19.8 (15.2) | 17.0 (14.2) | 19.2 (15.5) | 17.5 (13.5) |
| Home owned | 2403 (84.5%) | 1474 (86.2%) | 473 (79.1%) | 277 (85.8%) | 179 (84.0%) |
| Number of rooms (Mean (SD)) | 2.17 (1.23) | 2.19 (1.21) | 2.04 (1.11) | 2.28 (1.50) | 2.29 (1.22) |
| WATER, SANITATION AND HYGIENE (WASH) | | | | | |
| Improved drinking water source | 1667 (58.6%) | 1011 (59.1%) | 361 (60.4%) | 173 (53.6%) | 122 (57.3%) |
| Household treats drinking water | 643 (22.6%) | 392 (22.9%) | 128 (21.4%) | 79 (24.5%) | 44 (20.7%) |
| Improved sanitation facility in home | 1742 (61.3%) | 1070 (62.6%) | 329 (55.0%) | 209 (64.7%) | 134 (62.9%) |
| Basic hygiene facility in home | 311 (10.9%) | 197 (11.5%) | 45 (7.5%) | 36 (11.1%) | 33 (15.5%) |
| SPATIAL FACTORS | | | | | |
| Min. dist. (km) to drug shop (Mean (SD)) | 0.937 (1.15) | 0.967 (1.15) | 0.919 (1.20) | 0.925 (1.17) | 0.766 (0.984) |
| Min. dist. (km) to gov't. health centre (Mean (SD)) | 2.30 (1.33) | 2.34 (1.36) | 2.18 (1.22) | 2.22 (1.36) | 2.43 (1.29) |
| District | | | | | |
| Buliisa | 947 (33.3%) | 540 (31.6%) | 236 (39.5%) | 108 (33.4%) | 63 (29.6%) |
| Mayuge | 953 (33.5%) | 586 (34.3%) | 184 (30.8%) | 100 (31.0%) | 83 (39.0%) |
| Pakwach | 944 (33.2%) | 584 (34.2%) | 178 (29.8%) | 115 (35.6%) | 67 (31.5%) |

**Table S7.** Summary statistics of potential predictors for rejoining the study after an attrition event.

| | Total | Attriter | Rejoiner |
|---|---|---|---|
| | (N=1134) | (N=642) | (N=492) |
| BIOMEDICAL FACTORS | | | |
| Any symptoms | 44 (3.9%) | 23 (3.6%) | 21 (4.3%) |
| IDs for other HH members | 684 (60.3%) | 367 (57.2%) | 317 (64.4%) |
| NCDs for other HH members | 422 (37.2%) | 237 (36.9%) | 185 (37.6%) |
| History of IDs | 545 (48.1%) | 293 (45.6%) | 252 (51.2%) |
| History of NCDs | 193 (17.0%) | 92 (14.3%) | 101 (20.5%) |
| History of any disease | 617 (54.4%) | 331 (51.6%) | 286 (58.1%) |
| SOCIODEMOGAPHICS | | | |
| Age (Mean (SD)) | 22.6 (16.3) | 21.2 (15.4) | 24.4 (17.3) |
| Sex - Female | 631 (55.6%) | 354 (55.1%) | 277 (56.3%) |
| Tribe | | | |
| Alur | 630 (55.6%) | 327 (50.9%) | 303 (61.6%) |
| Bagungu | 108 (9.5%) | 57 (8.9%) | 51 (10.4%) |
| Musoga | 187 (16.5%) | 121 (18.8%) | 66 (13.4%) |
| Other | 209 (18.4%) | 137 (21.3%) | 72 (14.6%) |
| Majority religion | 808 (71.3%) | 454 (70.7%) | 354 (72.0%) |
| Highest level of education attained (Mean (SD)) | 3.72 (2.86) | 3.61 (2.82) | 3.88 (2.92) |
| Occupation | | | |
| Farmer | 172 (15.2%) | 92 (14.3%) | 80 (16.3%) |
| Fisherman | 98 (8.6%) | 43 (6.7%) | 55 (11.2%) |
| Fishmonger | 38 (3.4%) | 19 (3.0%) | 19 (3.9%) |
| None/Other | 826 (72.8%) | 488 (76.0%) | 338 (68.7%) |
| Home quality score (Mean (SD)) | 6.48 (3.64) | 6.64 (3.59) | 6.28 (3.69) |
| Household social status | 101 (8.9%) | 46 (7.2%) | 55 (11.2%) |
| Number of individuals in household (Mean (SD)) | 3.53 (1.24) | 3.51 (1.25) | 3.57 (1.24) |
| Deaths in household (past 3 yrs) | 60 (5.3%) | 41 (6.4%) | 19 (3.9%) |
| Years household has lived in village (Mean (SD)) | 17.7 (14.5) | 17.4 (14.6) | 18.1 (14.4) |
| Home owned | 929 (81.9%) | 520 (81.0%) | 409 (83.1%) |
| Number of rooms (Mean (SD)) | 2.15 (1.26) | 2.14 (1.22) | 2.16 (1.30) |
| WATER, SANITATION AND HYGIENE (WASH) | | | |
| Improved drinking water source | 656 (57.8%) | 361 (56.2%) | 295 (60.0%) |
| Household treats drinking water | 251 (22.1%) | 135 (21.0%) | 116 (23.6%) |
| Improved sanitation facility in home | 672 (59.3%) | 391 (60.9%) | 281 (57.1%) |
| Basic hygiene facility in home | 114 (10.1%) | 64 (10.0%) | 50 (10.2%) |
| SPATIAL FACTORS | | | |
| Min. dist. (km) to drug shop (Mean (SD)) | 0.892 (1.16) | 0.841 (1.12) | 0.958 (1.20) |
| Min. dist. (km) to gov't. health centre (Mean (SD)) | 2.24 (1.28) | 2.26 (1.28) | 2.21 (1.28) |
| District | | | |
| Buliisa | 407 (35.9%) | 222 (34.6%) | 185 (37.6%) |
| Mayuge | 367 (32.4%) | 237 (36.9%) | 130 (26.4%) |
| Pakwach | 360 (31.7%) | 183 (28.5%) | 177 (36.0%) |

**Table S8.** Attrition rates in study districts at major study timepoints.

| District | Jan-Feb 2023 | Oct 2023 | Jan-Feb 2024 |
|---|---|---|---|
| Pakwach | 18.9% (178/944) | 23.5% (222/944) | 20.6% (194/944) |
| Buliisa | 24.9% (236/947) | 26.3% (249/947) | 26.3% (249/947) |
| Mayuge | 19.3% (184/953) | 22% (210/953) | 27.6% (263/953) |

**Table S9.** Exposures and outcomes with later attrition.

| | Total | In | Out | P-value |
|---|---|---|---|---|
| | | Jan-Feb 2023 | | |
| | (N=2806) | (N=2215) | (N=591) | |
| S. mansoni (Jan-Feb 2022) | 1218 (43.4%) | 962 (43.4%) | 256 (43.3%) | 0.997 |
| Eggs per gram category (WHO) (Jan-Feb 2022) | | | | 0.978 |
| No | 1588 (56.6%) | 1253 (56.6%) | 335 (56.7%) | |
| High | 233 (8.3%) | 183 (8.3%) | 50 (8.5%) | |
| Mild | 361 (12.9%) | 288 (13.0%) | 73 (12.4%) | |
| Low | 624 (22.2%) | 491 (22.2%) | 133 (22.5%) | |
| Periportal fibrosis (Jan-Feb 2022) | 327 (11.7%) | 258 (11.6%) | 69 (11.7%) | 1 |
| Any schistosomiasis-related referrals (Jan-Feb 2022) | 73 (2.6%) | 60 (2.7%) | 13 (2.2%) | 0.585 |
| | | Oct 2023 | | |
| | (N=2207) | (N=1901) | (N=306) | |
| S. mansoni (Jan-Feb 2023) | 724 (32.8%) | 622 (32.7%) | 102 (33.3%) | 0.883 |
| Eggs per gram category (WHO) (Jan-Feb 2023) | | | | 0.526 |
| No | 1483 (67.2%) | 1279 (67.3%) | 204 (66.7%) | |
| High | 102 (4.6%) | 89 (4.7%) | 13 (4.2%) | |
| Mild | 165 (7.5%) | 136 (7.2%) | 29 (9.5%) | |
| Low | 457 (20.7%) | 397 (20.9%) | 60 (19.6%) | |
| Periportal fibrosis (Jan-Feb 2023) | 345 (15.6%) | 305 (16.0%) | 40 (13.1%) | 0.214 |
| Any schistosomiasis-related referrals (Jan-Feb 2023) | 5 (0.2%) | 5 (0.3%) | 0 (0%) | 1 |
| | | Jan-Feb 2024 | | |
| | (N=2135) | (N=1868) | (N=267) | |
| S. mansoni (Oct 2023) | 883 (41.4%) | 773 (41.4%) | 110 (41.2%) | 1 |
| Eggs per gram category (WHO) (Oct 2023) | | | | 0.999 |
| No | 1252 (58.6%) | 1095 (58.6%) | 157 (58.8%) | |
| High | 184 (8.6%) | 161 (8.6%) | 23 (8.6%) | |
| Mild | 220 (10.3%) | 192 (10.3%) | 28 (10.5%) | |
| Low | 479 (22.4%) | 420 (22.5%) | 59 (22.1%) | |
| | | Jan-Feb 2024 | | |
| | (N=2207) | (N=1855) | (N=352) | |
| S. mansoni (Jan-Feb 2023) | 724 (32.8%) | 592 (31.9%) | 132 (37.5%) | 0.047 |
| Eggs per gram category (WHO) (Jan-Feb 2023) | | | | 0.136 |
| No | 1483 (67.2%) | 1263 (68.1%) | 220 (62.5%) | |
| High | 102 (4.6%) | 87 (4.7%) | 15 (4.3%) | |
| Mild | 165 (7.5%) | 136 (7.3%) | 29 (8.2%) | |
| Low | 457 (20.7%) | 369 (19.9%) | 88 (25.0%) | |
| Periportal fibrosis (Jan-Feb 2023) | 345 (15.6%) | 297 (16.0%) | 48 (13.6%) | 0.296 |
| Any schistosomiasis-related referrals (Jan-Feb 2023) | 5 (0.2%) | 4 (0.2%) | 1 (0.3%) | 0.581 |

#### S3 Supplementary figures

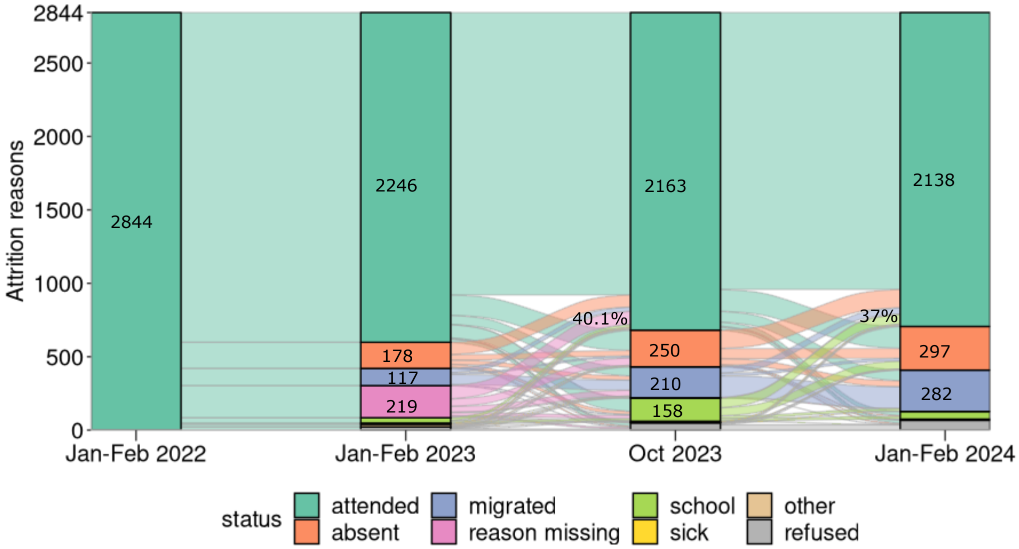


**Fig. S1.** Attrition reasons. Participant attendance across different timepoints, along with reasons for attrition.

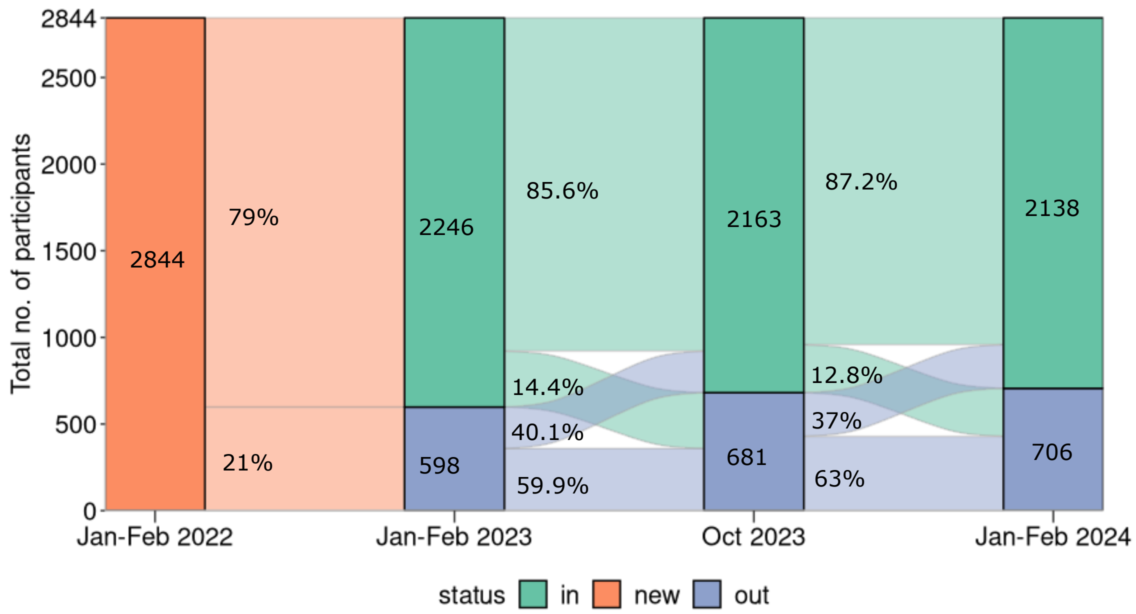


**Fig. S2.** Participant counts. Participant flows in and out of the study, with major follow-ups illustrated (Note: The March 2022 short-term follow-up for PZQ efficacy is shown in the Supplementary Figure S3)

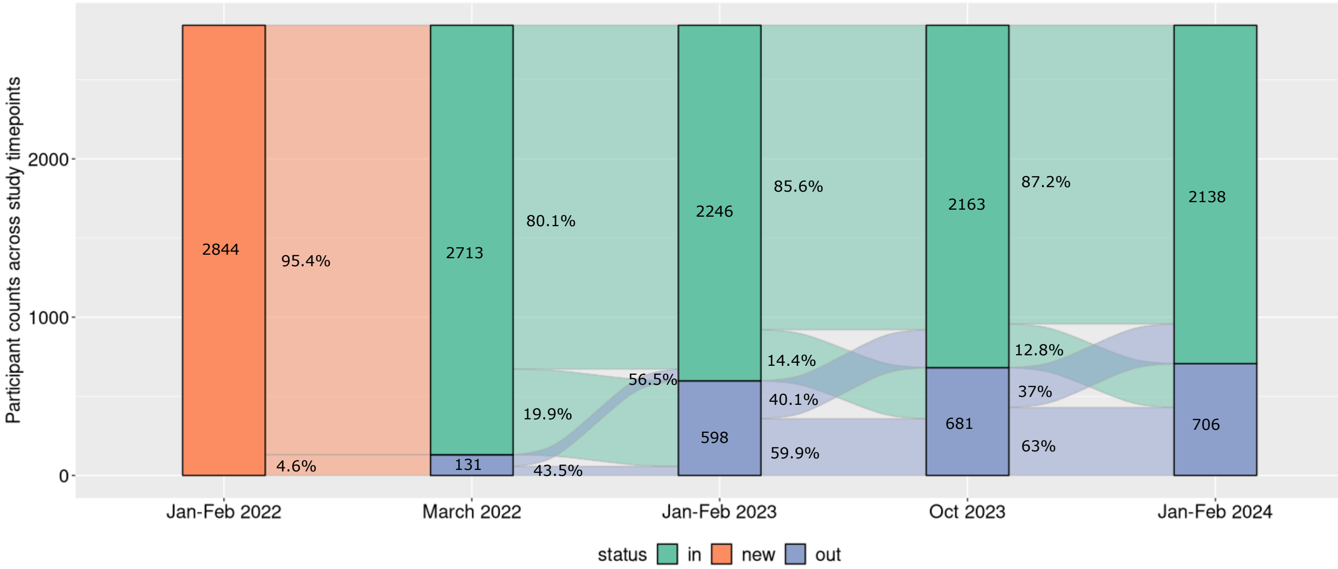


**Fig. S3.** Participant flows in and out of the study including March 2022 timepoint.

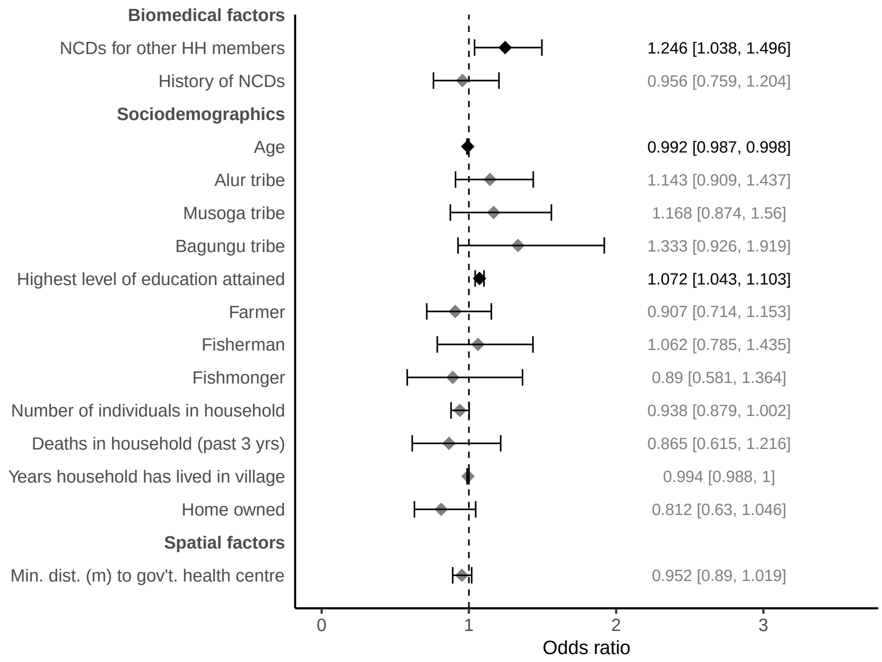


**Fig. S4.** Overall attrition model including short-term follow-up. Logistic regression model for overall attrition of study participants (n=2844) including the data collection timepoint of March 2022 with 95% confidence intervals calculated using household-level clustered standard errors (number of household clusters = 1445). VIFs < 10 for all variables. AUC for 10-fold cross-validation was 0.58. Results marked with a black diamond were significant and those marked in grey were non-significant.

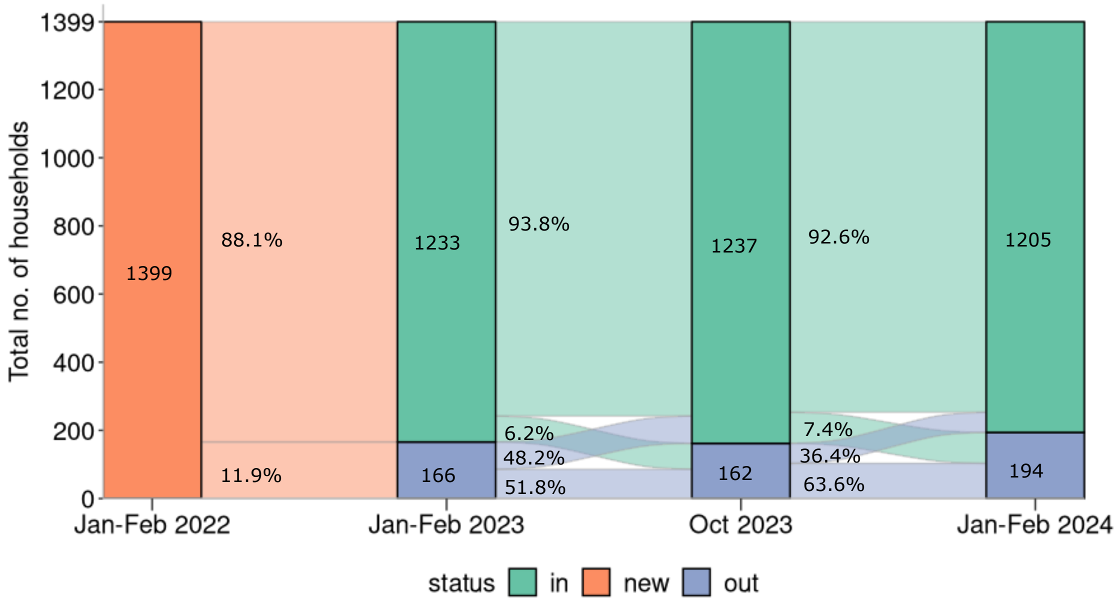


**Fig. S5.** Household counts. Households from which both participants missed a timepoint are labelled as out, while those from which at least one participant attended are labelled as in.

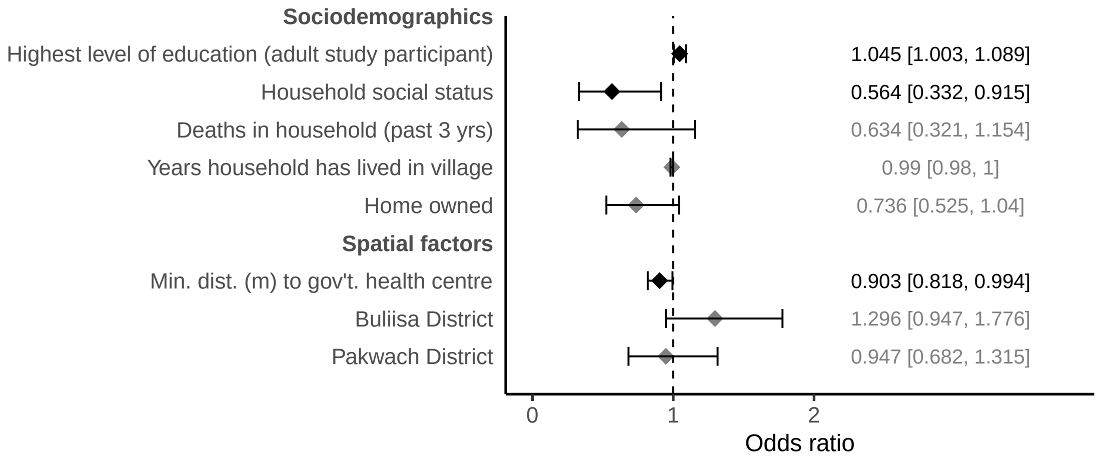


**Fig. S6.** Model for household attrition. Logistic regression model for household attrition (n=1399) with 95% confidence intervals. VIFs $< 10$ for all variables. AUC for 5-fold cross-validation was 0.6. 5-fold cross-validation was used due to the smaller sample size. Results marked with a black diamond were significant and those marked in grey were non-significant.

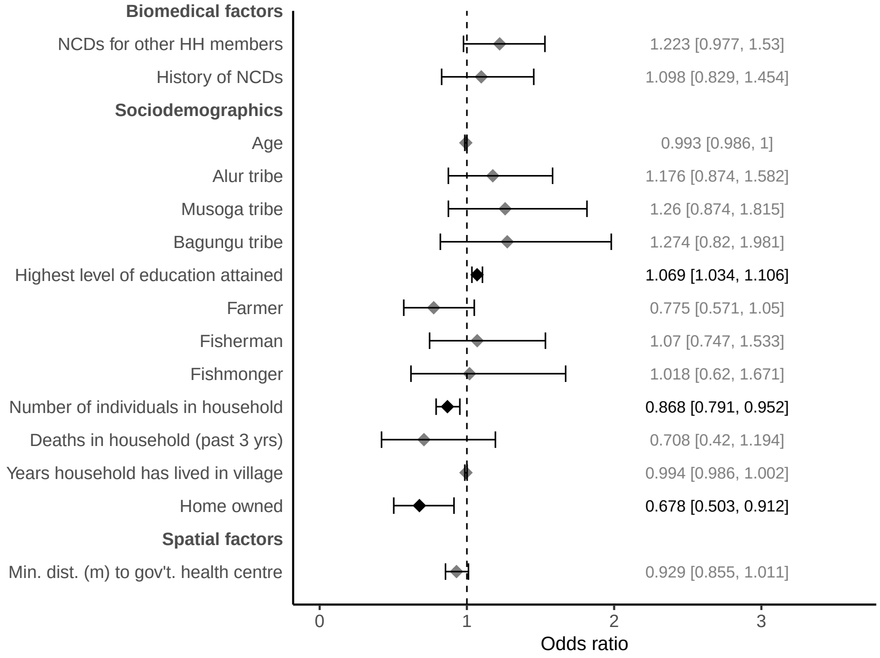


**Fig. S7.** Model for attrition in Jan-Feb 2023. Logistic regression model for attrition of study participants (n=2844) in Jan-Feb 2023 with 95% confidence intervals calculated using household-level clustered standard errors (number of household clusters = 1445). VIFs $< 10$ for all variables. AUC for 10-fold cross-validation was 0.58. Results marked with a black diamond were significant and those marked in grey were non-significant.

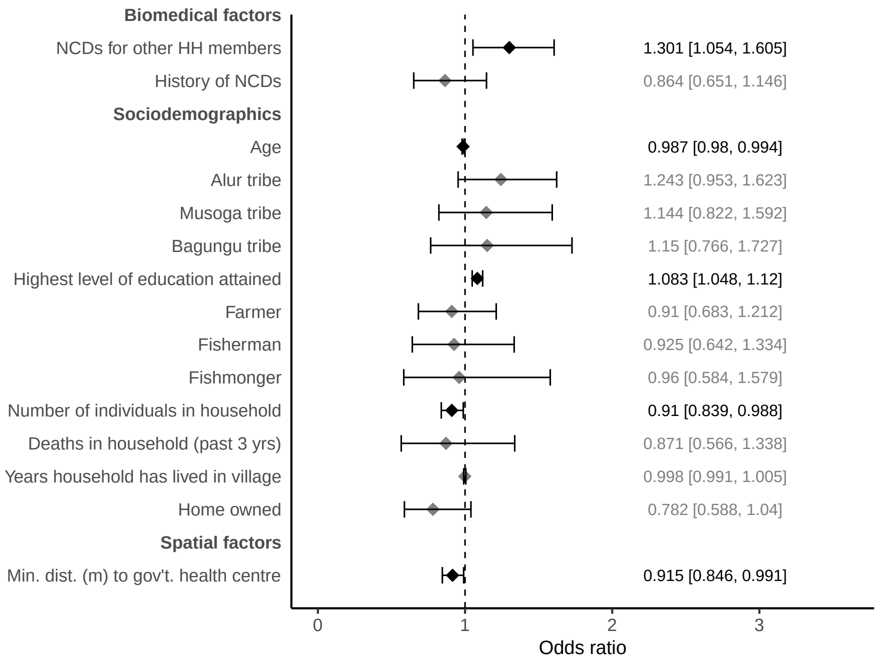


**Fig. S8.** Model for attrition in Oct 2023. Logistic regression model for attrition of study participants (n=2844) in Oct 2023 with 95% confidence intervals calculated using household-level clustered standard errors (number of household clusters = 1445). VIFs $< 10$ for all variables. AUC for 10-fold cross-validation was 0.6. Results marked with a black diamond were significant and those marked in grey were non-significant.

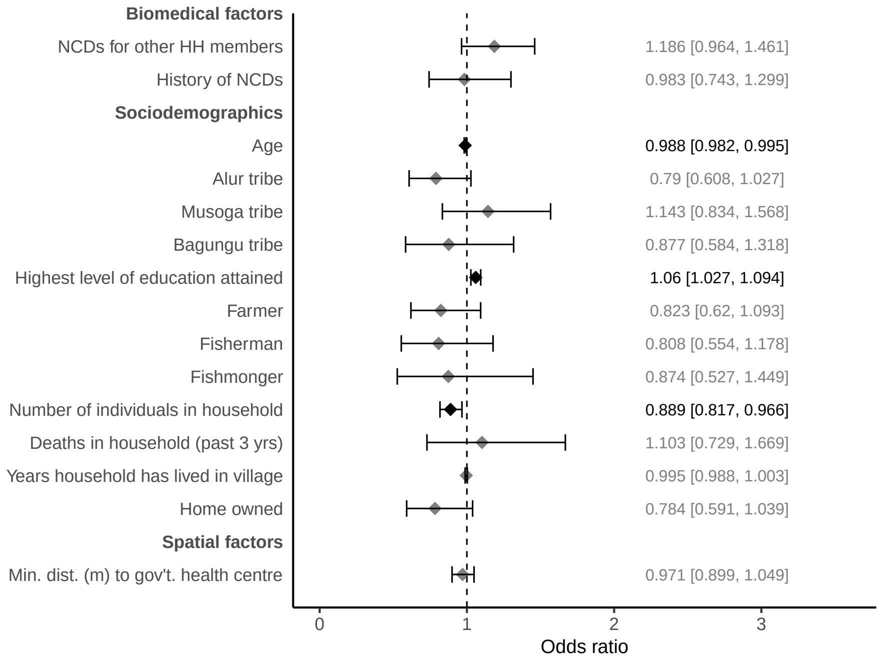


**Fig. S9.** Model for attrition in Jan-Feb 2024. Logistic regression model for attrition of study participants (n=2844) in Jan-Feb 2024 with 95% confidence intervals calculated using household-level clustered standard errors (number of household clusters = 1445). VIFs < 10 for all variables. AUC for 10-fold cross-validation was 0.59. Results marked with a black diamond were significant and those marked in grey were non-significant.

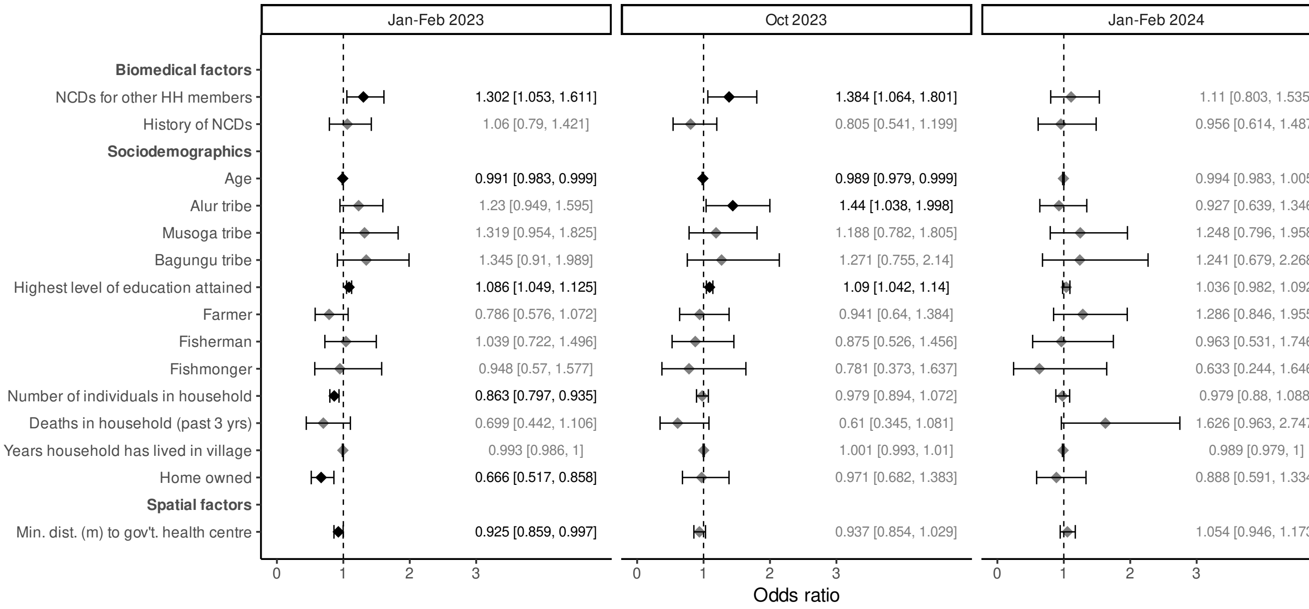


**Fig. S10.** Multinomial model for first attrition event. Multinomial regression model categorises the first attrition event of participants (n=2844) into one of three follow-up timepoints Jan-Feb 2023, Oct 2023, or Jan-Feb 2024, with not having an attrition event as the reference category. Odds ratios for predictor variables are shown with 95% confidence intervals. VIFs $< 10$ for all variables. AUC for 5-fold cross-validation was 0.57. 5-fold cross-validation was used due to the smaller sample sizes in individual categories. Results marked with a black diamond were significant and those marked in grey were non-significant.

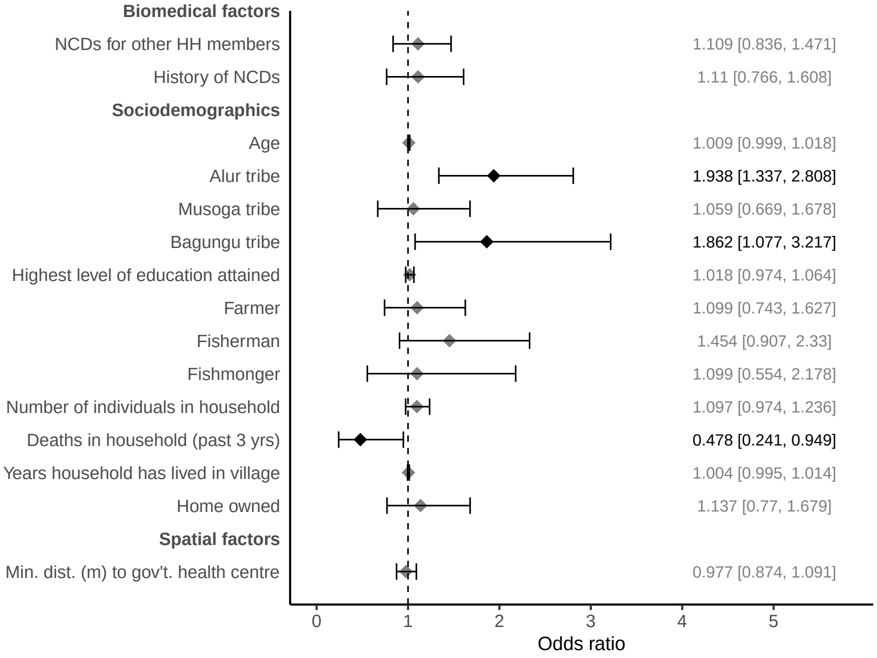


**Fig. S11.** Overall rejoiner model. Logistic regression model for rejoiners (n=1134) with 95% confidence intervals calculated using household-level clustered standard errors (number of household clusters = 795). VIFs < 10 for all variables. AUC for 5-fold cross-validation was 0.59. 5-fold cross-validation was used due to the smaller sample size. Results marked with a black diamond were significant, and those marked in grey were non-significant. Participants from the Alur tribe had significantly higher odds of attrition, with a FAR of 1.938 (95% CI: 1.634, 2.299) compared to the reference category. Similarly, participants from the Bagungu tribe also exhibited significantly higher attrition odds, with a FAR of 1.862 (95% CI: 1.256, 2.759). No significant difference was found for the Musoga tribe compared to the reference category.

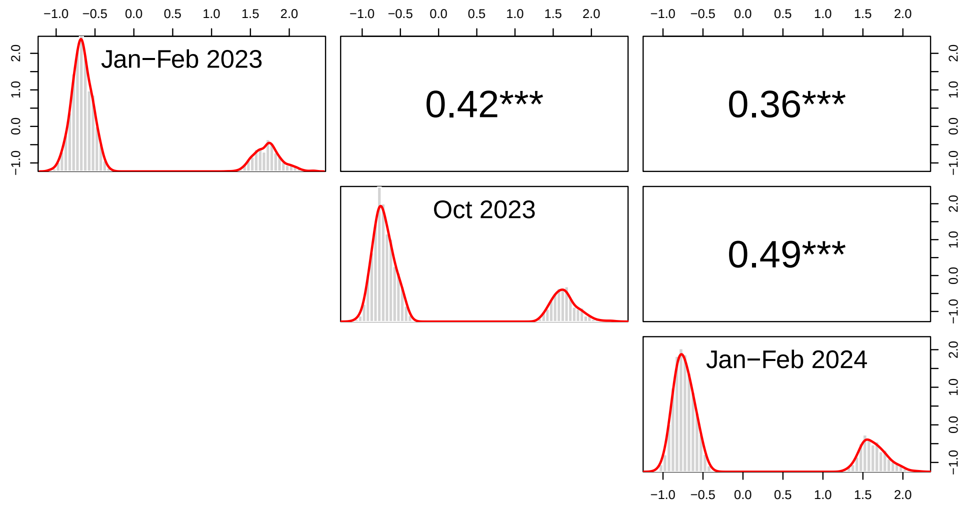


**Fig. S12.** Correlation of residuals. Correlations between residuals of logistic regression models of the major study time points.

#### References


1. Chami GF, Kontoleon AA, Bulte E, et al. Profiling nonrecipients of mass drug administration for schistosomiasis and hookworm infections: a comprehensive analysis of praziquantel and albendazole coverage in community-directed treatment in Uganda. Clinical Infectious Diseases. 2016;62(2):200-7.
2. World Health Organization. Improved sanitation facilities and drinking water sources;. [Accessed 6 Aug 2024]. Available: `https://www.who.int/data/nutrition/nlis/info/improved-sanitation-facilities-and-drinking-water-sources`.
3. World Health Organization, et al. Guidelines for drinking-water quality: incorporating the first and second addenda. World Health Organization; 2022.
4. World Health Organization. STEPwise approach to NCD risk factor surveillance (STEPS);. [Accessed 20 Aug 2024]. Available: `https://www.who.int/teams/noncommunicable-diseases/surveillance/systems-tools/steps`.
5. Hijmans RJ, Van Etten J, Mattiuzzi M, et al.. Raster package in R;. [Accessed 20 Aug 2024]. Available: `https://rspatial.org/raster/pkg/RasterPackage.pdf`.
6. Richter J, Hatz C, Campagne G, et al.. Ultrasound in schistosomiasis: a practical guide to the standardized use of ultrasonography for the assessment of schistosomiasis-related morbidity: second international workshop held in Niamey, Niger, 22-26 October, 1996.;.
7. Anjorin S, Nabatte B, Mpooya S, et al. The epidemiology of periportal fibrosis and relevance of current Schistosoma mansoni infection: a population-based, cross-sectional study. medRxiv. 2023:2023-09.
8. Katz N, Chaves A, Pellegrino J. A simple device for quantitative stool thick-smear technique in schistosomiasis mansoni. Revista do instituto de medicina tropical de São Paulo. 1972;14(6):397-400.
9. World Health Organization, et al. WHO guideline on control and elimination of human schistosomiasis. World Health Organization; 2022.
